# Multi-Trait Polygenic Scores for COPD and COPD Exacerbations Implicate Druggable Proteins

**DOI:** 10.1101/2025.08.22.25334001

**Authors:** Chengyue Zhang, Iain R. Konigsberg, Yixuan He, Jingzhou Zhang, Tinashe Chikowore, William B. Feldman, Xiaowei Hu, Yi Ding, Bogdan Pasaniuc, Diana Chang, Qingwen Chen, Jessica A. Lasky-Su, Julian Hecker, Martin D. Tobin, Jing Chen, Sean Kalra, Katherine A. Pratte, Hae Kyung Im, Emily S. Wan, Ani Manichaikul, Edwin K. Silverman, Russell P. Bowler, Leslie A. Lange, Victor E. Ortega, Alicia R. Martin, Michael H. Cho, Matthew R. Moll

## Abstract

**Objectives.:** To construct multi-trait polygenic scores (PRS) predicting chronic obstructive pulmonary disease (COPD) and exacerbations, validate their performance in diverse cohorts, and identify PRS-related proteins for potential therapeutic targeting.

**Design:** Prospective cohort studies.

**Setting.:** Genetic Epidemiology of COPD (COPDGene; 2007-present), Evaluation of COPD Longitudinally to Identify Predictive Surrogate Endpoints (ECLIPSE; 2005-2008), Mass General Brigham Biobank (MGBB; 2010-present), All of Us (2016-present), and UK Biobank (UKB; 2006-present).

**Participants.:** 6,647 non-Hispanic White (NHW) and 2,466 African American (AA) participants from COPDGene; 1,858 participants from ECLIPSE; 118,566 from All of Us; 15,142 from MGBB with genetic data. 5,173 COPDGene and 5,012 UKB participants with proteomic data.

**Main outcome measures.:** COPD status (GOLD 2-4 vs. GOLD 0) and COPD exacerbation frequency.

**Results.:** PRSmix+, a multi-trait PRS framework, selected 7 traits for a composite PRS (PRS_multi_). In multivariable models, PRS_multi_ was associated with COPD status (meta-analysis random effects (RE) OR 1.58 [95% CI: 1.28-1.94]) and exacerbation frequency (meta-analysis RE beta 0.21 [95% CI: 0.11-0.31]), with higher effect sizes observed in smoking-enriched cohorts. PRS_multi_ outperformed traditional single-trait PRS in all tested cohorts. Using protein prediction models, we identified 73 proteins associated with the PRS that were also validated with measured protein levels in COPDGene and UK biobank. Of these proteins, 25 were linked to approved or investigational drugs. Notable targets include AGER (RAGE), IL1RL1, and SCARF2, all implicated in COPD pathogenesis and exacerbations.

**Conclusions.:** Multi-trait PRS improves prediction of COPD and exacerbation risk. Integration with proteomic data identifies druggable protein targets, offering a promising avenue for precision medicine in COPD management.

**Trial registration.:** COPDGene: NCT00608764; ECLIPSE: NCT00292552.

## INTRODUCTION

Chronic obstructive pulmonary disease (COPD), a leading global cause of morbidity and mortality, is marked by persistent airflow limitation and an enhanced inflammatory lung response to harmful inhaled particles or gases. COPD is highly heterogeneous in presentation, progression, therapy response, and exacerbations, the latter of which are major drivers of morbidity^1^. Comorbid conditions, such as cardiovascular and metabolic diseases, are important predictors of COPD-related mortality. However, susceptibility to COPD and its comorbidities is determined by complex gene-by-environment interactions^2^. Notably, only a minority of smokers develop COPD, with genetics accounting for ∼30-40% of the variability in susceptibility^3^.

Genome-wide association studies (GWAS) have identified numerous genetic variants linked to lung function and COPD^4^. Summing GWAS variants together, polygenic risk scores (PRS) for two measures of lung function (forced expiratory volume at one second: FEV_1_ and the ratio of FEV_1_ to forced vital capacity: FEV_1_/FVC) were combined into a COPD PRS that was highly predictive of COPD in multiple cohorts, outperforming single PRSs in predicting COPD and COPD-related phenotypes^5^. Recent findings highlight that BMI genetics can predict mortality in COPD patients, underscoring the importance of multiple trait (multi-trait) genetic analyses in understanding COPD risk and outcomes^6^.

Multi-trait genetic analyses, which identify shared mechanisms among traits, have gained traction with the rise of a multitude of PRSs for various traits. He et al. developed a multi-trait PRS for COPD that performed exceptionally well in biobanks^7^. Further, this multi-trait PRS and the prior lung function-based COPD PRS were associated with exacerbations. However, in the latter, the COPD PRS effects were attenuated when accounting for baseline lung function, consistent with the known clinical association between COPD severity and exacerbations. It remains unclear whether combining PRSs for spirometry and broader phenotypes can better predict COPD and exacerbations across diverse cohorts.

While shared genetics can inform phenotype prediction and reveal insights into disease mechanisms, proteins offer several advantages. First, proteins are influenced by both genetics and environmental factors, such as infections - the leading cause of exacerbations^8^. Second, proteins can serve as direct therapeutic targets for small molecules^9^. Third, recent advances in statistical tools can use genetics to predict protein expression levels^10^ which are more likely causally related to a trait than non-genetically regulated proteins^11^. We hypothesized that integrating PRSs for spirometry and comorbid traits could enhance the prediction of COPD and exacerbations in both research and biobank cohorts. Furthermore, we sought to identify and validate genetically predicted protein levels linked to shared genetic architectures, using measured protein levels to confirm key findings.

## METHODS

### Study Cohorts

#### COPDGene

The Genetic Epidemiology of COPD (COPDGene) study (ClinicalTrials.gov Identifier: NCT00608764) is a smoking-enriched cohort of self-identified non-Hispanic White (NHW) and African American (AA) participants aged 45-80 years with ≥ 10 pack-years of smoking history^12^. COPDGene had enrollment, 5- and 10-year visits. Proteomic data were collected at visit 2 (5-year follow-up).

#### ECLIPSE

The Evaluation of COPD Longitudinally to Identify Predictive Surrogate Endpoints (ECLIPSE) study (ClinicalTrials.gov Identifier: NCT00292552) was a multicenter cohort study designed to explore biomarkers and clinical phenotypes of COPD^13^. Participants include individuals with moderate to severe COPD (GOLD stages 2-4), aged 40-75 years, recruited across 12 countries.

#### MGBB

The Mass General Brigham Biobank (MGBB) is a large-scale biorepository linked to electronic health records (EHRs) of over 117,000 participants from the Mass General Brigham healthcare system since 2009. Participants aged ≥18 years were genotyped using the Illumina Global Screening Array.

#### All of Us

The All of Us Research Program is a population-scale biobank designed to advance precision medicine by collecting genetic, environmental, and health data from over one million participants in the United States. Whole-genome sequencing was conducted using the Illumina NovaSeq platform as previously described^14^. We utilized the allele call/allele frequency (ACAF) dataset for analyses. Additional cohort details, genotyping information, and proteomic details are in the

## Supplementary Appendix

### Statistical analysis

#### Overview of study design

We integrated genetic data from COPDGene, ECLIPSE, MGBB, UKB, and All of Us to develop and validate a multi-trait PRS (PRS_multi_) for both COPD and exacerbations. Following genotype imputation and PRS construction, we applied PRSmix+ to combine trait-specific PRSs relevant to COPD susceptibility and exacerbation risk^15^. We assessed associations with COPD case-control status and exacerbations. In COPDGene, NHW and AA participants were analyzed separately due to differences in demographic and disease characteristics, partly shaped by recruitment strategies. In other cohorts, racial/ethnic groups were analyzed together, adjusting for self-identified race and principal components of genetic ancestry.

To identify underlying biological mechanisms, we integrated genetically predicted protein levels (via S-PrediXcan) with measured plasma proteomic data from COPDGene and UKBB to identify proteins linked to COPD and exacerbations^10^. These candidate proteins were then cross-referenced with drug databases to highlight potential targets for therapeutic repurposing. An overview of the study design is shown in **Figure 1**.

**Figure 1.**
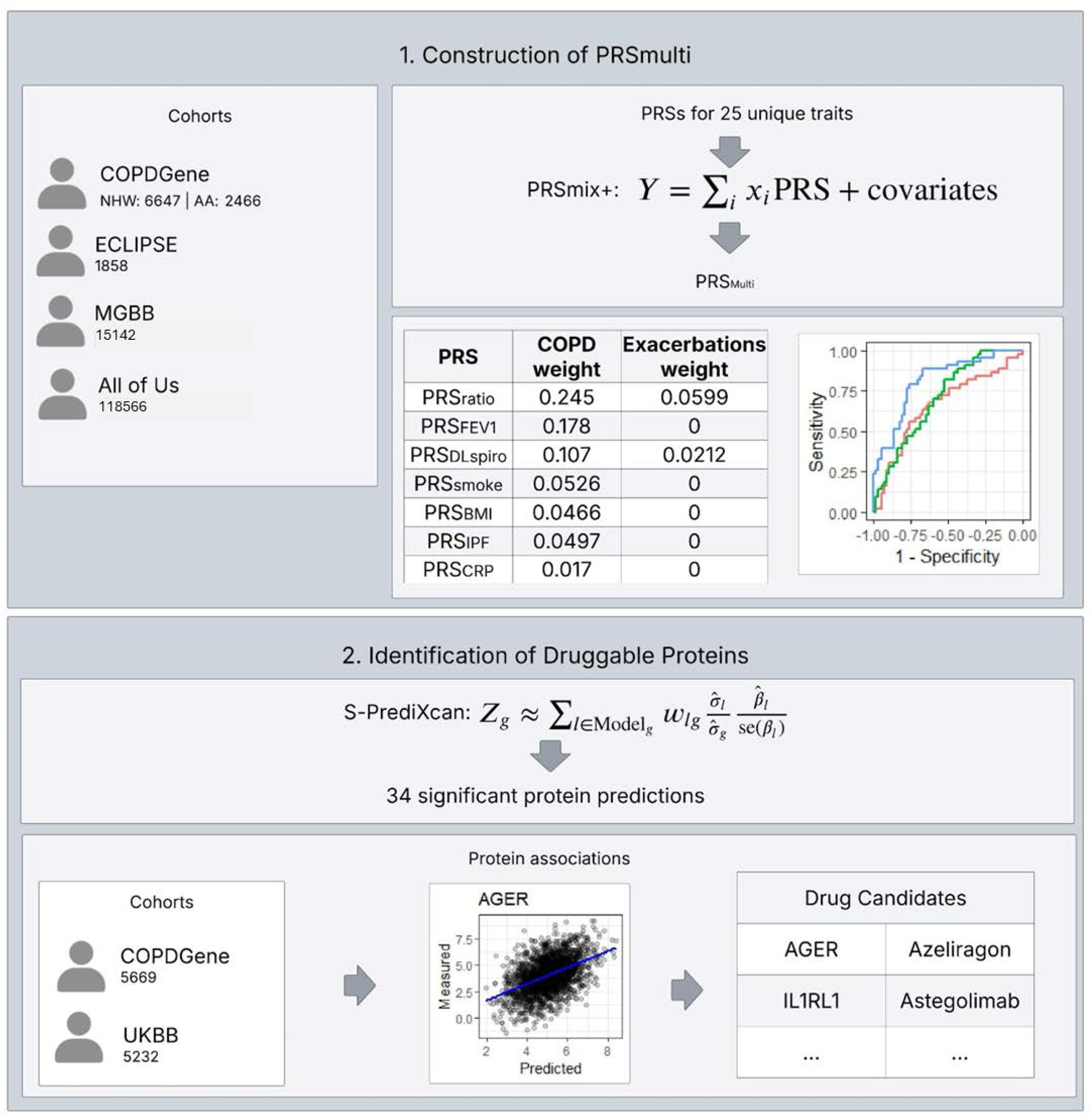
Overview of Study Design. Schematic representation of the study workflow. Plots included in this figure are intended for illustrative purposes only.

## Outcomes

### COPD

#### COPD in Research Cohorts

COPD was defined using the GOLD (Global Initiative for Chronic Obstructive Lung Disease) classification system. Two groups were identified: GOLD 2-4, representing those with moderate to very severe airflow limitation (FEV_1_/FVC<0.7 and FEV_1_ < 80% predicted), and GOLD 0, referring to those with normal spirometry (FEV_1_/FVC≥0.7 and FEV_1_ ≥ 80% predicted).

#### COPD in Biobanks

For genetic analyses, we included individuals aged ≥40 years with available smoking data who were ever-smokers. We excluded participants with other lung diseases aside from asthma (based on ICD-9-CM codes 516.x, 515.x, 428.x and ICD-10-CM codes J84.x, I50.x). The index date was defined as the date of the first COPD exacerbation or the first recorded observation for controls. COPD was defined by ≥1 inpatient code or ≥3 outpatient diagnostic codes within the past three years^16^ to ensure identification of individuals with active disease. We identified COPD patients using ICD-9-CM codes 491.xx, 492.xx, 496 and ICD-10-CM codes J41.x, J42, J43.x, J44.x^17^.

In UKBB, COPD case-control status was defined using spirometry to align with the definition in research cohorts (see below).

### COPD Exacerbations

#### Exacerbations in Research Cohorts

In COPDGene, annualized exacerbation rates were calculated from the first study visit using data from the Longitudinal Follow-up Program, which was designed to continuously collect data on clinical outcomes, including exacerbations, comorbidities, and mortality, through surveys and electronic health records^18^. As COPDGene was enriched for individuals who smoke, and individuals without COPD in this cohort experience respiratory events and increased mortality, we included these individuals in analyses^19^. In ECLIPSE, exacerbation data were collected at each study visit using standardized questionnaires that recorded the number of exacerbations and use of steroids or antibiotics since the prior visit. As a result of study inclusion and exclusion criteria, nearly all participants with genetic data had had moderate-to-severe COPD.

In COPDGene, we additionally assessed quantitative computed tomography (CT) measures of emphysema (volume-noise-bias-adjusted lung density^20^) and airway pathology (Pi10: square root of the wall area of a hypothetical airway with an internal perimeter of 10 millimeters).

#### Exacerbations in Biobanks

COPD exacerbation numbers were tracked for one year following the index date. Events were identified using ICD-9-CM codes 491.21, 493.22 and ICD-10-CM codes J44.0, J44.1. Multiple events within a 15-day period were considered a single exacerbation. For proteomic analysis, only exacerbations occurring after proteomic sampling were included in the analysis.

### Predictors

#### Single-trait polygenic risk scores

PRSs were computed using GWAS summary statistics or obtained from the PGS Catalog^21,22^. Selection of PRSs was based on biological relevance according to clinician input and literature review, available summary statistics, and the exclusion of testing cohorts from the original GWAS to avoid overfitting.

Details on PRS development are provided in the **Supplementary Appendix**, and variant weights are available upon request.

#### Development of a multi-trait polygenic risk score

Prior to constructing a multi-trait PRS, we oriented directions of effects of individual PRSs such that a higher PRS is associated with higher COPD risk (e.g. higher PRS_FEV1_ is associated with higher COPD risk) since it is not clear what the expected effects are for all traits. Several methods exist for calculating multi-trait PRSs. A detailed description of PRSmix and PRSmix+^15^ is in the **Supplementary Appendix**.

We trained PRSmix+ using COPDGene non-Hispanic white participants, as most component PRSs were originally developed in populations with high genetic similarity to European reference panels. Models were trained to predict COPD, COPD exacerbations, Pi10, and emphysema, adjusting for age, sex, smoking pack-years, BMI, five genetic principal components, and CT scanner model (as appropriate). We selected the model that incorporated the largest number of PRSs and used the resulting weights to construct the final multi-trait PRS (PRS_multi_). All PRSs were centered and scaled prior to analysis, ensuring a mean of 0 and a standard deviation (SD) of 1.

#### Testing of the multi-trait polygenic risk score

We tested the association of PRS_multi_ with COPD and exacerbations using multivariable logistic regression models and negative binomial models, respectively. All models were adjusted for age, sex, smoking pack-years, BMI, and five genetic principal components. Exacerbation models included a log-offset for follow-up time. For comparison, we performed these same regression analyses using each individual component PRS included in PRS_multi_. To account for multiple comparisons, statistical significance was defined as a Benjamini-Hochberg (BH) adjusted p-value less than 0.05. As a sensitivity analysis, we further adjusted models for baseline FEV_1_ in research cohorts. We performed area under the receiver operating characteristic curve (AUC) analyses, which are detailed in the **Supplementary Appendix**.

#### Meta-analysis

Following multivariable regression analyses, we performed random effects meta-analyses using the meta R package for PRS_multi_ and component PRSs^23^. Heterogeneity was assessed using the I² statistic, and funnel plots were generated to visualize inter-cohort variability and assess selection bias.

#### Protein prediction

We applied the S-PrediXcan framework, which leverages GWAS summary statistics to infer associations between predicted protein expression and complex traits or diseases, to evaluate genetically predicted protein levels. Summary statistics from the PRSs contributing to PRS_multi_^10^ served as inputs. S-PrediXcan combines protein-wide association models with GWAS summary statistics, enabling the identification of protein-trait associations without requiring individual-level data. Prior to analysis, we harmonized GWAS variants to the GTEx v8 reference panel^24^ and imputed summary statistics to address ambiguous or missing variants. We then applied multi-ancestry protein prediction models from the Atherosclerosis Risk in Communities (ARIC) study^25^ to estimate genetically regulated proteins with significantly altered levels.

For significant proteins (BH FDR-adjusted *p*-value < 0.1), we assessed whether directly measured protein levels were associated with COPD status and exacerbations in both COPDGene and UK Biobank. In COPDGene, models were adjusted for age, sex, race, pack-years of smoking, COPD case-control status, and log offset of time. In UK Biobank, analyses used propensity score matching on these variables, except the log offset of time since exacerbations were followed for 1 year, to account for case-control imbalance.

#### Drug repurposing analysis

Based on the above analyses, we identified a list of targetable proteins that satisfied the following criteria: 1) significantly predicted to have altered levels based on GWAS/S-PrediXcan results, 2) significant association with COPD exacerbations, 3) concordant direction of effect between UK Biobank and COPDGene. From the resulting list of proteins, we further prioritized proteins by the number of PRS_multi_-associated traits for which all three criteria were met. See the Supplementary Appendix for further details.

## RESULTS

### Characteristics of study participants

A total of 144,679 individuals were included in genetic analyses (**Table 1**), drawn from both COPD-enriched cohorts (COPDGene and ECLIPSE) and population-scale biobanks (MGBB and All of Us). As expected, smoking prevalence varied across cohorts.

**Table 1.** Characteristics of study participants. Baseline demographic and clinical characteristics of participants across cohorts (COPDGene, ECLIPSE, MGBB, UKBB, and All of Us). COPDGene = Genetic Epidemiology of COPD. ECLIPSE = Evaluation of COPD Longitudinally to Identify Predictive Surrogate Endpoints. MGBB = MassGeneral Brigham Biobank. NHW = non-Hispanic white. AA = African American.

| *Characteristic* | *COPDGene NHW* | *COPDGene AA* | *ECLIPSE* | *All of Us* | *MGBB* |
| --- | --- | --- | --- | --- | --- |
| N | 6647 | 2466 | 1858 | 118566 | 15142 |
| Age in years (mean (sd)) | 62.03 (8.83) | 54.90 (7.44) | 63.14 (7.52) | 60.71 (11.37) | 64.22 (12.08) |
| Sex (No. % Female) | 3170 (47.7) | 1045 (42.4) | 625 (33.6) | 71119 (60.0) | 8918 (58.9) |
| smoking status (No. %) |  |  |  |  |  |
| Never | 0 | 0 | 0 | 75915 (64.9) | 8489 (56.1) |
| Former | 4045 (60.9) | 529 (21.5) | 1217 (65.5) | 22650 (19.4) | 5835 (38.5) |
| Current | 2602 (39.1) | 1937 (78.5) | 641 (34.5) | 18486 (15.8) | 818 (5.4) |
| BMI (mean (sd)) | 28.69 (6.04) | 28.76 (6.48) | 26.77 (5.53) | 30.12 (7.30) | 27.84 (6.02) |
| Pack years of smoking (mean (sd)) | 47.26 (26.02) | 38.47 (21.05) | 48.92 (27.75) | 6.79 (15.37) |  |
| FEV1 % predicted (mean (sd)) | 77.85 (27.49) | 74.15 (23.19) | 52.20 (22.53) |  |  |
| FEV1/FVC (mean (sd)) | 0.64 (0.17) | 0.71 (0.15) | 0.47 (0.15) |  |  |
| COPD (No. % Cases) | 2634 (48.6) | 803 (40.6) | 1711 (92.1) | 3980 (3.4) | 459 (3.0) |
| Exacerbation rate (No./year) | 0.12 (0.51) | 0.14 (0.66) | 0.75 (0.98) | 0.04 (0.29) | 0.01 (0.17) |

Cohort characteristics for COPDGene and UKBB proteomic analyses are shown in **Table S1**. Participants had similar age, sex distribution, and spirometric measures. In COPDGene, individuals who were never smokers were excluded.

### Development of multi-trait polygenic risk scores for COPD and related traits

We used 25 individual PRSs (**Table S2**), chosen based on clinician input and literature review, to develop multi-trait PRSs for COPD and related traits. Four new PRSs were developed for this analysis (emphysema, peak expiratory flow, eosinophils, and smoking) which are available upon request. Clustering analysis revealed that PRSs for spirometry were highly correlated and that the emphysema and Type 2 diabetes scores were correlated, as were asthma/allergic rhinitis and venous thromboembolism/cor pulmonale (**Figure S1**).

We constructed multi-trait PRS models for COPD status (GOLD 2-4 vs. GOLD 0) and exacerbations, as well as CT measures of quantitative emphysema (adjusted lung density) and airway wall thickness (Pi10) using PRSmix+. Mixing weights for each model are displayed in **Table S3**. Final weighted models included between two and seven individual PRSs. Notably, PRSs for FEV_1_/FVC (PRS_ratio_) and a deep learning spirometry (PRS_DLspiro_) trait contribute to all 4 traits. PRSs for FEV_1_ (PRS_FEV1_), BMI (PRS_BMI_), and CRP (PRS_CRP_) contribute to both Pi10 and COPD, and all seven PRSs, including smoking (cigarettes per day) (PRS_smoke_) and idiopathic pulmonary fibrosis (PRS_IPF_), contribute to COPD. Thus, while PRS_ratio_ is the largest contributor to COPD genetic risk, six additional PRSs also contribute to COPD risk.

All multi-trait PRSs showed strong associations with their respective training outcomes in multivariable models (**Table S4**). As our a priori decision was to select the multi-trait PRS that represents the highest number of comorbid traits, the PRS for COPD derived from 7 separate PRSs was carried forward for further testing and will hereafter be referred to as the PRS_multi_. The PRS_multi_ explained 20.7% of the variance on the liability scale for COPD^26^.

### Testing the multi-trait polygenic risk score for COPD

We tested the predictive utility of PRS_multi_ for both COPD and COPD exacerbations compared with individual component PRSs. For COPD, PRS_multi_ consistently demonstrated larger absolute effect sizes in all cohorts except All of Us, where PRS_BMI_ showed the strongest effect (**Table S5**). For COPD exacerbations, PRS_multi_ outperformed other scores in COPDGene NHW and ECLIPSE, whereas PRS_BMI_ had higher effects in the remaining cohorts (**Table S6**). Notably, PRS_multi_ associations with COPD status remained significant after adjusting for baseline FEV_1_ in COPDGene, but associations with exacerbations were attenuated in FEV_1_-adjusted models.

In a random effects meta-analysis, PRS_multi_ was associated with increased odds of COPD (OR: 1.58 per SD (95% confidence interval (CI): 1.28 - 1.94)) (**Figure 2A**). We observed significant between-study heterogeneity (I^2^ = 0.98, *p* < 0.01), with stronger effects in smoking-enriched research cohorts. Among the component PRSs, PRS_IPF_ and PRS_BMI_ were the only scores not significantly associated with COPD, and PRS_multi_ had a larger effect on COPD status than any individual score (**Figure 2B**).

**Figure 2.**
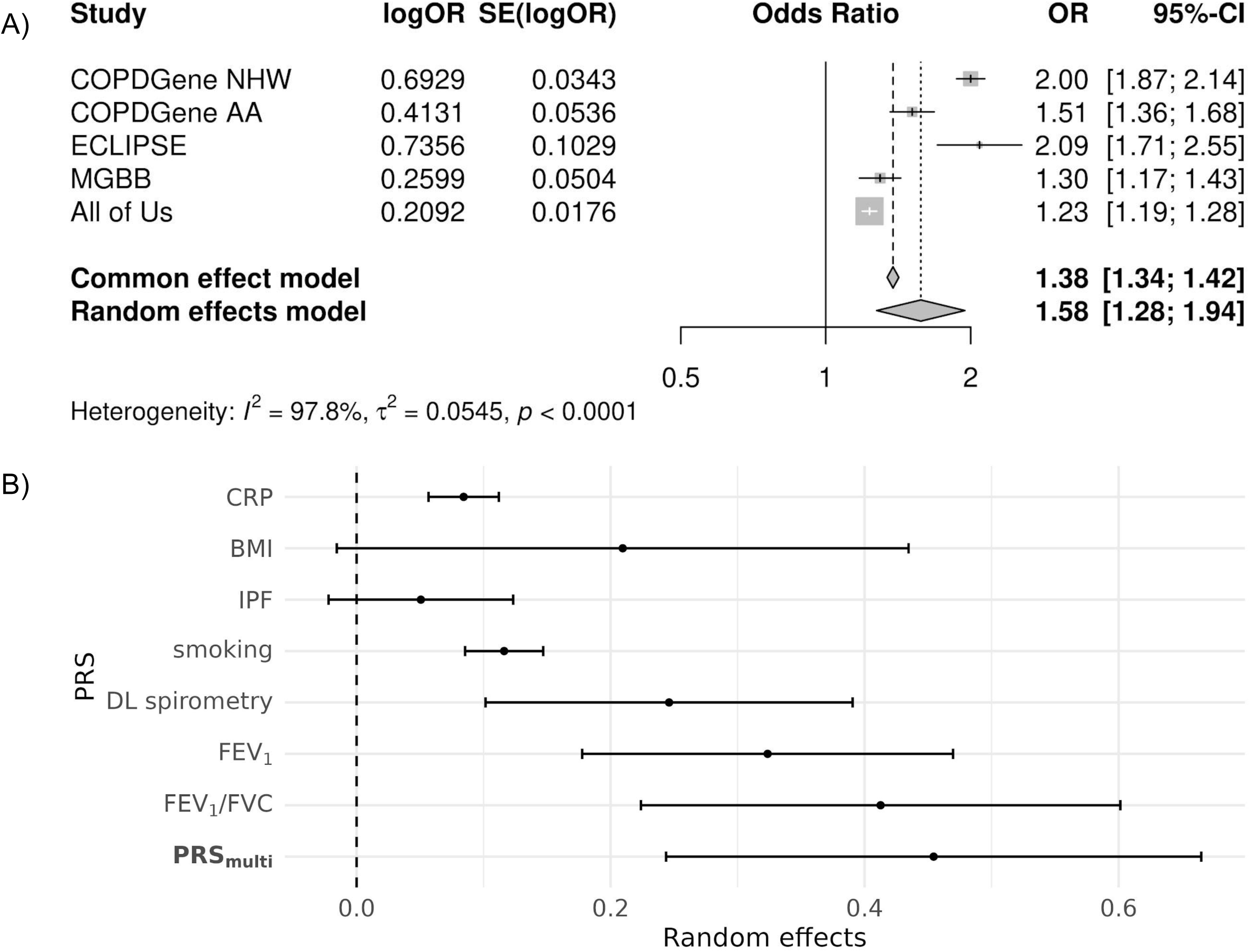
PRS_multi_ is associated with COPD. **A.** Forest plot from a meta-analysis of the association between PRS_multi_ and COPD status across multiple cohorts. The figure presents odds ratios (ORs) with 95% confidence intervals (CIs) for each cohort, demonstrating the overall effect size and heterogeneity (I²). **B.** Forest plot comparing the effects of individual PRSs and PRS_multi_ on COPD risk. PRS_multi_ outperforms single-trait PRSs, particularly in smoking-enriched cohorts. CRP = C-reactive protein. BMI=body-mass index. IPF=idiopathic pulmonary fibrosis. DL=deep learning. PRS=polygenic risk score. Cohort abbreviations are in the legend for Table 1.

We next performed a similar analysis for COPD exacerbations but with multivariable negative binomial modeling of count data. In meta-analysis, PRS_multi_ was associated with an increase of 0.21 exacerbations per year per SD (95% CI: 0.11 - 0.31) (**Figure 3A**). However, this association was attenuated in sensitivity analyses adjusting for baseline FEV_1_. All component PRSs were significantly associated with exacerbations except for the PRS_IPF_ and PRS_BMI_ (**Figure 3B**). We note that PRS_BMI_ demonstrated the largest effects on exacerbations compared to other PRSs in some cohorts (**Table S6**), but it was not significantly associated with exacerbations in meta-analysis (**Figure S2**) due to a flipped direction of effect in COPDGene AA participants. Again, between-study heterogeneity was significant (I^2^ = 0.86, *p* < 0.0001), though less pronounced than for COPD status.

**Figure 3.**
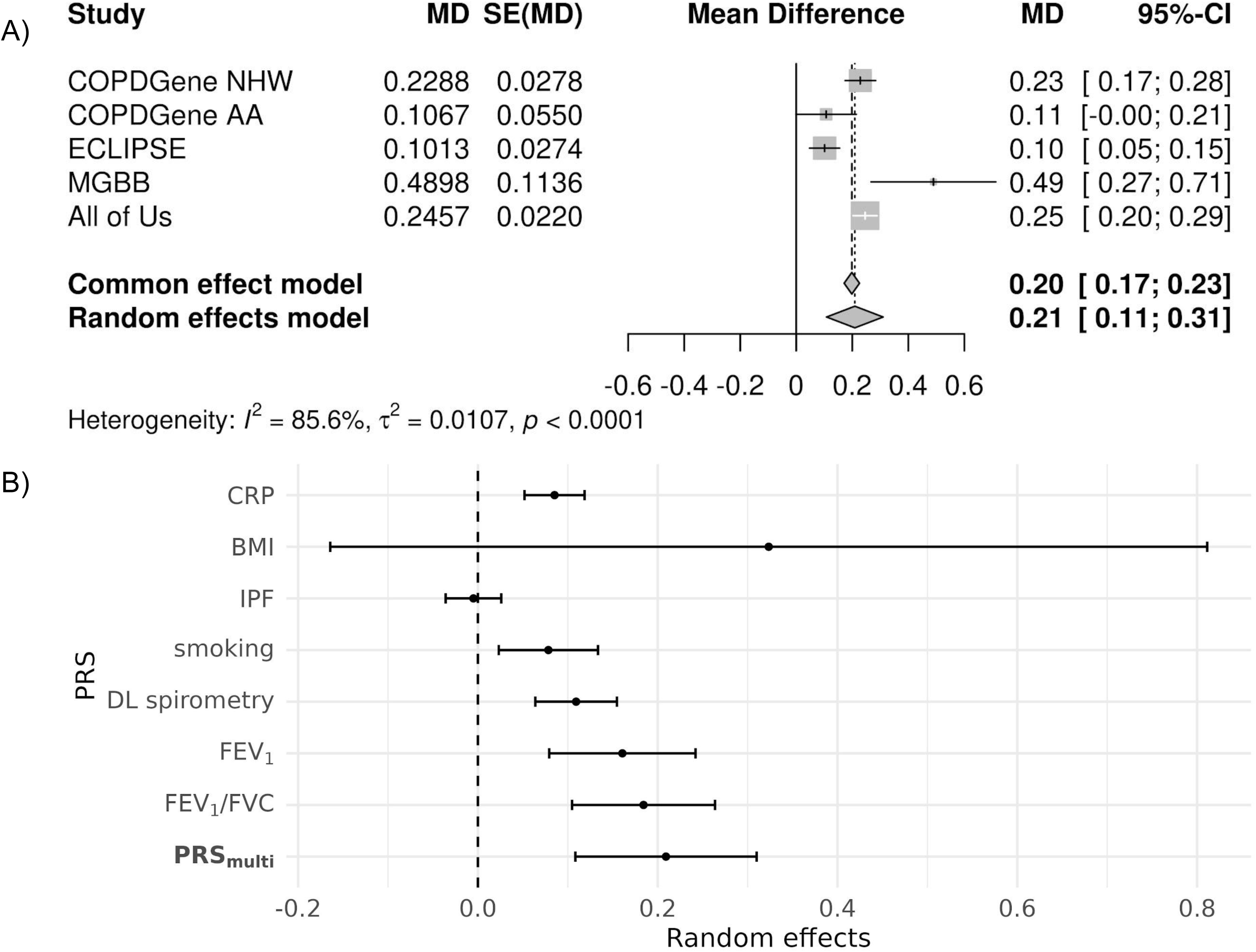
PRS is associated with COPD exacerbations. **A.** Meta-analysis forest plot showing the association between PRS_multi_ and COPD exacerbations, with estimates from individual cohorts and the overall effect. The PRS_multi_ is significantly associated with increased exacerbation risk. **B.** Comparison of individual PRSs and PRS_multi_ for exacerbation prediction. Effect sizes and confidence intervals are presented, highlighting PRS_multi_ as the most predictive score. Abbreviations are in the legends of Figure 2 and Table 1.

We also observed that heterogeneity in COPD status associations was driven largely by differences between research and biobank cohorts, reflecting the use of spirometry-versus ICD-based case definitions, respectively (**Figure S3, top**). In contrast, heterogeneity in exacerbation outcomes was lower and appeared to be influenced by outlier results from MGBB participants (**Figure S3, bottom**). In a leave-one-out sensitivity analysis excluding COPDGene NHW participants, we observed similar results for COPD and COPD exacerbations (**Figure S4**)

In AUC analyses among COPDGene NHW participants, PRS_multi_ outperformed PRS_FEV1/FVC_ in predicting both COPD (AUC 0.67 vs 0.64, p = 1.05 x 10^-8^) and frequent exacerbations (≥ 2 per year) (AUC 0.569 vs 0.562, p = 0.000001). Both PRSs improve prediction when added to clinical covariates (**Table 2** & **Figure S5**).

**Table 2.**
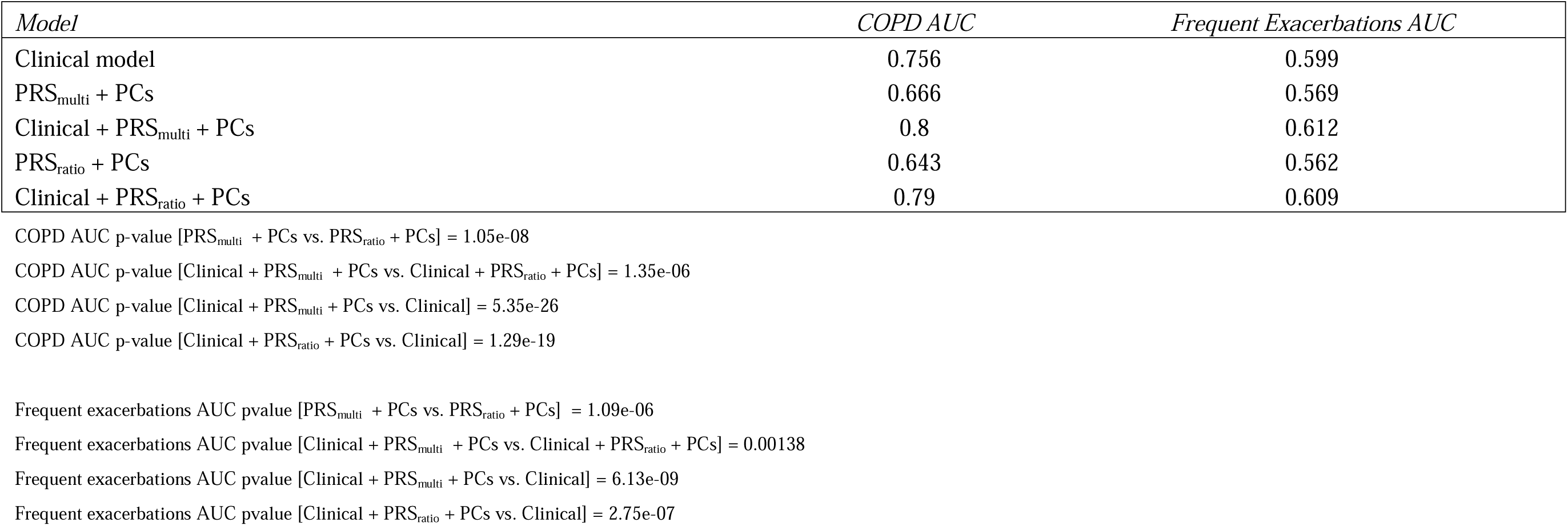
Area-under-the-curve (AUC) analyses comparing PRS_multi_ to single PRSs for COPD and exacerbations. AUC comparisons for PRS_multi_ versus the best-performing single PRSs for predicting COPD and frequent exacerbations (≥2 per year) in COPDGene NHW participants. The table also reports AUC changes when adding PRS to a clinical model (age, sex, BMI, smoking pack-years), with DeLong test p-values for model comparison.

| *Model* | *COPD AUC* | *Frequent Exacerbations AUC* |
| --- | --- | --- |
| Clinical model | 0.756 | 0.599 |
| PRS<sub>multi</sub> + PCs | 0.666 | 0.569 |
| Clinical + PRS<sub>multi</sub> + PCs | 0.8 | 0.612 |
| PRS<sub>ratio</sub> + PCs | 0.643 | 0.562 |
| Clinical + PRS<sub>ratio</sub> + PCs | 0.79 | 0.609 |

COPD AUC p-value [PRS<sub>multi</sub> + PCs vs. PRS<sub>ratio</sub> + PCs] = 1.05e-08
COPD AUC p-value [Clinical + PRS<sub>multi</sub> + PCs vs. Clinical + PRS<sub>ratio</sub> + PCs] = 1.35e-06
COPD AUC p-value [Clinical + PRS<sub>multi</sub> + PCs vs. Clinical] = 5.35e-26
COPD AUC p-value [Clinical + PRS<sub>ratio</sub> + PCs vs. Clinical] = 1.29e-19
Frequent exacerbations AUC pvalue [PRS<sub>multi</sub> + PCs vs. PRS<sub>ratio</sub> + PCs] = 1.09e-06
Frequent exacerbations AUC pvalue [Clinical + PRS<sub>multi</sub> + PCs vs. Clinical + PRS<sub>ratio</sub> + PCs] = 0.00138
Frequent exacerbations AUC pvalue [Clinical + PRS<sub>multi</sub> + PCs vs. Clinical] = 6.13e-09
Frequent exacerbations AUC pvalue [Clinical + PRS<sub>ratio</sub> + PCs vs. Clinical] = 2.75e-07

### Genetically-predicted proteins based on shared genetic risks amongst traits

Following the identification of seven traits contributing to COPD genetic risk, we investigated shared genetic mechanisms by identifying proteins whose levels are predicted to be altered based on the genetic architecture of these traits. Using GWAS summary statistics for each trait, we applied S-PrediXcan^10^ with ARIC PredictDB protein expression models^25^ to infer genetically regulated protein levels (full results available upon request). We then tested whether measured plasma protein levels were associated with COPD exacerbations using multivariable negative binomial regression models in COPDGene and UKBB. Models were adjusted for COPD case-control status and other confounders (see **Methods**). We examined only proteins meeting FDR-adjusted significance in S-PrediXcan, COPDGene SomaScan (measured protein levels), and UKBB Olink (measured protein levels) data (**Table S7**). We further restricted the protein list to those with concordant directions of effects in COPDGene and UKBB, resulting in a final set of 73 genetically predicted proteins associated with COPD exacerbations (**Table 3**).

**Table 3.**
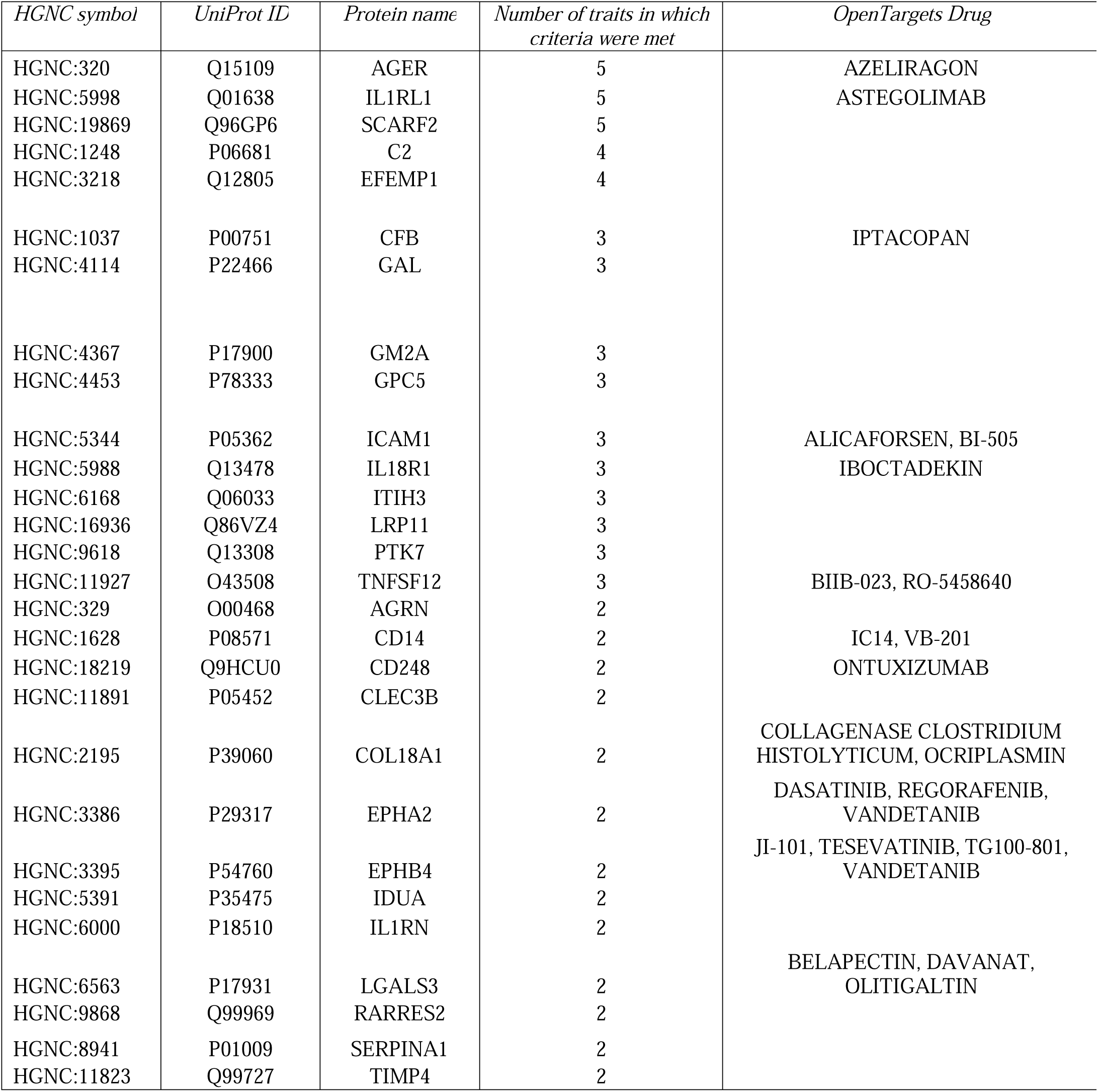
Druggable proteins associated with COPD exacerbations. List of targetable proteins satisfying three criteria: (1) significant genetic association via S-PrediXcan, (2) measured protein level significantly associated with exacerbations in UKBB and COPDGene, and (3) concordant effect directions in both cohorts. Prioritization is based on the number of PRS_multi_-associated traits involved. The table also includes drug targets from OpenTargets. Only proteins for which 2 or more traits were implicated are shown.

We ranked these proteins by the number of traits for which all criteria were met. The advanced glycosylation end-product specific receptor (AGER), interleukin 1 receptor-like 1 (IL1RL1), and scavenger receptor class F member 2 (SCARF2) were represented in 5 out of 7 tested traits. SERPINA1 was represented in two out of 7 tested traits. We performed gene name lookups in the OpenTargets Platform^27^ to identify potential drug repurposing candidates. We identified 25 proteins with existing clinical trials for 46 drugs (**Table 3 and Table S8**). Notably, we found drugs targeting AGER and IL1RL1 that are being tested in clinical trials, with the latter being tested in COPD. Finally, among the 73 proteins, we identified eight proteins (Agrin (AGRN), CD300C, CFB, GM2 Ganglioside Activator (GM2A), IL1RL1, INHBB, Leukocyte Immunoglobulin Like Receptor A5 (LILRA5), TIMP4) that demonstrated consistent associations with COPD status, frequent exacerbations, and the highest vs. lowest quintile of PRS_multi_ in COPDGene NHW participants (**Figure S6**).

## DISCUSSION

In this study, we leveraged genetic data from over 140,000 individuals across smoking-enriched research cohorts and large-scale biobanks to develop a multi-trait polygenic risk score (PRS_multi_) for COPD and exacerbations. PRS_multi_ outperformed single-trait PRSs in predicting both COPD risk and exacerbation frequency. Using elastic net modeling, we developed a multi-trait PRS and identified seven component traits that share a genetic basis with COPD, including spirometry measures such as FEV_1_/FVC and FEV_1_, a deep learning-derived spirometry phenotype, cigarettes per day, pulmonary fibrosis, C-reactive protein (CRP), and BMI. Using the genetic associations for these traits, we applied protein prediction models and identified 73 genetically predicted proteins whose measured levels were associated with exacerbations in both a smoking-enriched and a biobank cohort, independent of COPD status. These findings provide critical insights into the shared genetic architecture of COPD and related traits and nominate a set of proteins as potential biomarkers and therapeutic targets.

Although COPD comorbidities are known predictors of exacerbations and mortality, their genetic contributions to COPD risk have been underexplored. We identified seven traits contributing to the genetic risk of COPD, emphasizing its complex genetic architecture. Among these, three are spirometry-based, including a deep learning-derived spirometry phenotype, indicating that traditional measures like FEV_1_ and FEV_1_/FVC alone may not fully capture genetic risk or phenotypic variability. Notably, the PRSs of several traits known to be associated with exacerbations were not significant in elastic net modeling, including eosinophils and asthma, which have previously demonstrated genetic overlap with COPD^28,29^. This observation could reflect inclusion of relevant overlapping loci into other risk scores (e.g. asthma risk loci included in COPD or lung function), phenotypic specificity (e.g. data on eosinophilic COPD was not uniformly available), or other factors. These findings emphasize the need to understand the shared genetic architecture of COPD and inflammation-related traits.

To investigate shared genetic mechanisms between these seven traits, we used genetic protein prediction models derived from protein quantitative trait loci from the Atherosclerosis Risk in Communities (ARIC) study^25^. Using genetics to predict other omics has been shown to enhance prediction of complex traits; for example, polygenic transcriptome risk scores for spirometry using PrediXcan demonstrated greater portability across self-identified race and ethnicity groups compared to a standard PRS^30^.

Instead of focusing on a single trait, we applied a genetic protein prediction approach across seven traits and validated our findings using measured protein levels in two cohorts. As a result, we identified 73 genetically-predicted proteins associated with exacerbations in both cohorts after adjusting for COPD case-control status. To find the most relevant potential biomarkers for exacerbations, we analyzed proteins that were elevated in COPD cases, frequent exacerbators (≥2 exacerbations per year), and individuals in the highest quintile of PRS_multi_; we identified eight key proteins: AGRN, CD300C, CFB, GM2A, IL1RL1, INHBB, LILRA5, and TIMP4.

These findings suggest that PRS_multi_ and these proteins could be measured concurrently to predict COPD exacerbation risk. Developing an integrated prediction model and subsequent prospective validation are needed As drugs with genetically-backed targets lead to approval rates twice that of non-genetically-backed compounds, and drug repurposing agents achieve markedly higher approval rates (30% compared to 10% for de novo drugs), we used the genetically-predicted proteins associated with exacerbations to identify drug repurposing candidates^31–33^. Our analysis revealed 25 proteins with trials involving 46 drugs.

Based on 5 out of 7 traits, three proteins were genetically predicted to affect COPD and exacerbation risk: IL1RL1, AGER, and SCARF2. IL1RL1 is targeted by astegolimab, which is already in Phase 3 trials for COPD. This finding offers a proof-of-concept that our method can highlight potential drug repurposing candidates for COPD exacerbations. AGER (also known as s-RAGE), a highly replicable GWAS locus and well-established COPD biomarker^34^, is expressed in alveolar epithelial cells and appears to have a broad role in regulating immunity and inflammation. As such, AGER has been implicated in several diseases, including glioblastoma multiforme (NCT05986851), triple-negative breast cancer^35^, and Alzheimer’s disease^36^. Azeliragon is an antagonist of AGER and was well tolerated in a phase III trial of Alzheimer’s disease, but the trial (NCT02080364) was terminated due to lack of efficacy. The complicated issue in using azeliragon in COPD is that rs2070600 C➔T variant, associated with higher COPD risk, is associated with lower soluble RAGE (sRAGE) levels, and lower sRAGE levels are associated with more emphysema^37^. The effects of this variant on exacerbations are less clear, and further investigation needs to be done on which patients might benefit from repurposing azeliragon for COPD exacerbations.

SCARF2 polymorphisms have been implicated in COPD risk based on Mendelian randomization^38^, and we now report that this genetically predicted protein is associated with exacerbations at the measured protein level in two cohorts. We did not identify any drug repurposing candidates in OpenTargets based on SCARF2, making this protein a potential novel therapeutic target for exacerbations.

Strengths of this study are that it combines many participants from both research and biobank cohorts with carefully defined COPD and COPD exacerbation criteria. The use of PRSmix+ facilitated the identification of key traits contributing to genetic risk and the discovery of genetically predicted protein biomarkers, which were validated in two cohorts with measured protein levels. Additionally, the study identified several promising biomarkers and therapeutic targets based on genetic evidence.

There are several limitations. PRSmix+ studies typically aim to improve prediction, and while including more PRSs as inputs may enhance predictive power, we are approaching the upper bound of heritability for COPD, limiting further gains. The list of traits analyzed is not exhaustive, and translating findings from European to non-European cohorts remains a challenge. Cohort design limitations, such as recruiting only non-Hispanic white and African American individuals in COPDGene, may also influence results. However, we found similar results in All of Us when including all groups, emphasizing the value of diverse representative cohorts. Importantly, PRS_multi_ associations with exacerbations were attenuated when adjusting for baseline FEV_1_ in research cohorts, suggesting the effects are primarily driven by disease severity; by contrast, measured protein associations were adjusted for COPD status, suggesting that these genetically predicted protein targets are also important for disease activity and exacerbation risk. Finally, the integration of genetic and protein biomarkers, coupled with prospective validation and clinical implementation studies, is crucial to establish the practical utility and clinical relevance of these findings.

In conclusion, multi-trait PRSs for COPD and exacerbations identify genetic contributions to disease heterogeneity and druggable protein targets. These findings suggest that genetic risk prediction can be linked to specific therapeutic strategies for COPD precision medicine.

---

**Supplementary Figure 1.** Heatmap showing hierarchical clustering of Pearson correlation coefficients between PRSs input into PRSmix+.

**Supplementary Figure 2.** Forest plot showing the random effects meta-analysis of multivariable negative binomial associations of PRS_BMI_ on exacerbations.

**Supplementary Figure 3.** Funnel plots of meta-analysis results for COPD and exacerbations. Funnel plots displaying meta-analysis results for (A) COPD and (B) COPD exacerbations.

**Supplementary Figure 4.** Leave-one-out meta-analysis for COPD (A) and exacerbations **(B).** Meta-analyses excluding COPDGene NHW participants were performed, as this cohort was used to tune PRS_multi_ weights and could be prone to overfitting.

**Supplementary Figure 5. ROC curves for PRS-based models of COPD and exacerbations.** Receiver operating characteristic (ROC) curves for models predicting (A) COPD and (B) exacerbations. Models include combinations of clinical covariates, PRS_multi_, PRS_ratio_, and genetic principal components.

**Supplementary Figure 6.** Protein Associations with COPD Outcomes. Of the 73 genetically-predicted proteins identified to be associated with COPD exacerbations, eight proteins were associated with COPD affection status (left), frequent exacerbations (≥ 2 exacerbations/year vs. no exacerbations) (middle), and significantly differentially expressed between the top and bottom quintile of the PRS_multi_ (right) in COPDGene NHW participants. Boxplots for each association with interquartile ranges are displayed. P-values for Student t-tests are also displayed.

**Supplementary Table 1. Characteristics of study participants with proteomics data.** Summary statistics for participants with available plasma proteomics data from COPDGene and UKBB.

**Supplementary Table 2. PRSs included in PRSmix+ analysis.** List of PRSs used in PRSmix+ modeling, along with their source (GWAS, PGS Catalog, or newly developed) and references.

**Supplementary Table 3. PRS-mix weights for COPDGene, weighted by different phenotypes.** Summary of PRS-mix weights assigned to different traits in the construction of PRS_multi_. This table presents the contribution of each PRS to the final model predicting COPD and exacerbations.

**Supplementary Table 4. Association of PRSmix+ values with different weights in COPDGene.** Results of multivariable models testing PRSmix+ scores with different weights for their association with COPD and exacerbations.

**Supplementary Table 5. Multivariable associations of PRSs in PRSmix+ with COPD in each cohort.** Logistic regression results for PRSs in PRSmix+ predicting COPD across individual cohorts. Blue shading indicates the highest performing PRS in a cohort.

**Supplementary Table 6. Multivariable associations of PRSs in PRSmix+ with exacerbations in each cohort.** Negative binomial regression results for PRSs in PRSmix+ predicting exacerbation frequency across individual cohorts. Blue shading indicates the highest performing PRS in a cohort.

**Supplementary Table 7. S-PrediXcan proteins significantly associated with COPD and exacerbations.** List of proteins with significant associations based on S-PrediXcan using GWAS summary statistics for PRS_multi_ traits that were also significant in measuring proteomic association analysis using COPDGene SomaScan and UKB Olink data.

**Supplementary Table 8. Clinical trial information from OpenTargets for drugs targeting S-PrediXcan proteins.** Summary of drugs targeting proteins identified via S-PrediXcan analysis. Includes drug names, indications, and clinical trial phase information.

## Data Availability

Some of the data produced in the present work are contained in the manuscript. Further data are available upon reasonable request to the authors.

## Notes

*Funding*: MM is supported by NIH grant K08HL159318. EKS is supported by NIH grants R01 HL152728, P01 HL114501, U01 HL089856, R01 HL147148, and R01 HL133135. MHC is supported by NIH grants R01 HL153248, R01 HL135142, R01 HL149861, R01 HL137927, and R01 HL137148. COPDGene is also supported by the COPD Foundation through contributions made to an Industry Advisory Board that has included AstraZeneca, Bayer Pharmaceuticals, Boehringer-Ingelheim, Genentech, GlaxoSmithKline, Novartis, Pfizer, and Sunovion.

### Competing Interest Statement

MM discloses consulting fees from 2ndMD, TheaHealth, Axon Advisors, Dialectica, Sanofi, Verona Pharma; grant support from Genentech; service as a medical expert; and honoraria from ATS 2024 and NYSTS 2024.

### Funding Statement

Matthew R. Moll is supported by NIH grant K08HL159318. Edwin K. Silverman is supported by NIH grants R01 HL152728, P01 HL114501, U01 HL089856, R01 HL147148, and R01 HL133135. Michael H. Cho is supported by NIH grants R01 HL153248, R01 HL135142, R01 HL149861, R01 HL137927, and R01 HL137148. COPDGene is also supported by the COPD Foundation through contributions made to an Industry Advisory Board that has included AstraZeneca, Bayer Pharmaceuticals, Boehringer-Ingelheim, Genentech, GlaxoSmithKline, Novartis, Pfizer, and Sunovion.

### Author Declarations

Ethics committee/IRB of Mass General Brigham gave ethical approval for this work.

