## Supplementary Appendix for "Multi-Trait Polygenic Scores for COPD and COPD Exacerbations Implicate Druggable Proteins"

### Funding and Acknowledgements

**COPDGene Phase 3 Grant Support and Disclaimer**

The project described was supported by Award Number U01 HL089897 and Award Number U01 HL089856 from the National Heart, Lung, and Blood Institute. The content is solely the responsibility of the authors and does not necessarily represent the official views of the National Heart, Lung, and Blood Institute or the National Institutes of Health.

**COPD Foundation Funding**

COPDGene is also supported by the COPD Foundation through contributions made to an Industry Advisory Board that has included AstraZeneca, Bayer Pharmaceuticals, Boehringer- Ingelheim, Genentech, GlaxoSmithKline, Novartis, Pfizer, and Sunovion.

**COPDGene® Investigators – Core Units**

*Administrative Center*: James D. Crapo, MD (PI); Edwin K. Silverman, MD, PhD (PI); Barry J. Make, MD; Elizabeth A. Regan, MD, PhD

*Genetic Analysis Center*: Terri H. Beaty, PhD; Peter J. Castaldi, MD, MSc; Michael H. Cho, MD, MPH; Dawn L. DeMeo, MD, MPH; Adel El Boueiz, MD, MMSc; Marilyn G. Foreman, MD, MS; Auyon Ghosh, MD; Lystra P. Hayden, MD, MMSc; Craig P. Hersh, MD, MPH; Jacqueline Hetmanski, MS; Brian D. Hobbs, MD, MMSc; John E. Hokanson, MPH, PhD; Wonji Kim, PhD; Nan Laird, PhD; Christoph Lange, PhD; Sharon M. Lutz, PhD; Merry-Lynn McDonald, PhD; Dmitry Prokopenko, PhD; Matthew Moll, MD, MPH; Jarrett Morrow, PhD; Dandi Qiao, PhD; Elizabeth A. Regan, MD, PhD; Aabida Saferali, PhD; Phuwanat Sakornsakolpat, MD; Edwin K. Silverman, MD, PhD; Emily S. Wan, MD; Jeong Yun, MD, MPH

*Imaging Center*: Juan Pablo Centeno; Jean-Paul Charbonnier, PhD; Harvey O. Coxson, PhD; Craig J. Galban, PhD; MeiLan K. Han, MD, MS; Eric A. Hoffman, Stephen Humphries, PhD; Francine L. Jacobson, MD, MPH; Philip F. Judy, PhD; Ella A. Kazerooni, MD; Alex Kluiber; David A. Lynch, MB; Pietro Nardelli, PhD; John D. Newell, Jr., MD; Aleena Notary; Andrea Oh, MD; Elizabeth A. Regan, MD, PhD; James C. Ross, PhD; Raul San Jose Estepar, PhD; Joyce Schroeder, MD; Jered Sieren; Berend C. Stoel, PhD; Juerg Tschirren, PhD; Edwin Van Beek, MD, PhD; Bram van Ginneken, PhD; Eva van Rikxoort, PhD; Gonzalo Vegas Sanchez- Ferrero, PhD; Lucas Veitel; George R. Washko, MD; Carla G. Wilson, MS;

*PFT QA Center, Salt Lake City, UT*: Robert Jensen, PhD

*Data Coordinating Center and Biostatistics*, *National Jewish Health, Denver, CO*: Douglas Everett, PhD; Jim Crooks, PhD; Katherine Pratte, PhD; Matt Strand, PhD; Carla G. Wilson, MS

*Epidemiology Core*, *University of Colorado Anschutz Medical Campus, Aurora, CO*: John E. Hokanson, MPH, PhD; Erin Austin, PhD; Gregory Kinney, MPH, PhD; Sharon M. Lutz, PhD; Kendra A. Young, PhD

*Mortality Adjudication Core:* Surya P. Bhatt, MD; Jessica Bon, MD; Alejandro A. Diaz, MD, MPH; MeiLan K. Han, MD, MS; Barry Make, MD; Susan Murray, ScD; Elizabeth Regan, MD; Xavier Soler, MD; Carla G. Wilson, MS

*Biomarker Core*: Russell P. Bowler, MD, PhD; Katerina Kechris, PhD; Farnoush Banaei- Kashani, PhD

**COPDGene® Investigators – Clinical Centers**

*Ann Arbor VA:* Jeffrey L. Curtis, MD; Perry G. Pernicano, MD

*Baylor College of Medicine, Houston, TX*: Nicola Hanania, MD, MS; Mustafa Atik, MD; Aladin Boriek, PhD; Kalpatha Guntupalli, MD; Elizabeth Guy, MD; Amit Parulekar, MD;

*Brigham and Women’s Hospital, Boston, MA*: Dawn L. DeMeo, MD, MPH; Craig Hersh, MD, MPH; Francine L. Jacobson, MD, MPH; George Washko, MD

*Columbia University, New York, NY*: R. Graham Barr, MD, DrPH; John Austin, MD; Belinda D’Souza, MD; Byron Thomashow, MD

*Duke University Medical Center, Durham, NC*: Neil MacIntyre, Jr., MD; H. Page McAdams, MD; Lacey Washington, MD

*HealthPartners Research Institute, Minneapolis, MN*: Charlene McEvoy, MD, MPH; Joseph Tashjian, MD

*Johns Hopkins University, Baltimore, MD*: Robert Wise, MD; Robert Brown, MD; Nadia N. Hansel, MD, MPH; Karen Horton, MD; Allison Lambert, MD, MHS; Nirupama Putcha, MD, MHS

*Lundquist Institute for Biomedical Innovation at Harbor UCLA Medical Center, Torrance, CA*: Richard Casaburi, PhD, MD; Alessandra Adami, PhD; Matthew Budoff, MD; Hans Fischer, MD; Janos Porszasz, MD, PhD; Harry Rossiter, PhD; William Stringer, MD

*Michael E. DeBakey VAMC, Houston*, *TX*: Amir Sharafkhaneh, MD, PhD; Charlie Lan, DO *Minneapolis VA:* Christine Wendt, MD; Brian Bell, MD; Ken M. Kunisaki, MD, MS

*Morehouse School of Medicine, Atlanta, GA*: Eric L. Flenaugh, MD; Hirut Gebrekristos, PhD; Mario Ponce, MD; Silanath Terpenning, MD; Gloria Westney, MD, MS

*National Jewish Health, Denver, CO*: Russell Bowler, MD, PhD; David A. Lynch, MB *Reliant Medical Group, Worcester, MA*: Richard Rosiello, MD; David Pace, MD

*Temple University, Philadelphia, PA:* Gerard Criner, MD; David Ciccolella, MD; Francis Cordova, MD; Chandra Dass, MD; Gilbert D’Alonzo, DO; Parag Desai, MD; Michael Jacobs, PharmD; Steven Kelsen, MD, PhD; Victor Kim, MD; A. James Mamary, MD; Nathaniel Marchetti, DO; Aditi Satti, MD; Kartik Shenoy, MD; Robert M. Steiner, MD; Alex Swift, MD; Irene Swift, MD; Maria Elena Vega-Sanchez, MD

*University of Alabama, Birmingham, AL:* Mark Dransfield, MD; William Bailey, MD; Surya P. Bhatt, MD; Anand Iyer, MD; Hrudaya Nath, MD; J. Michael Wells, MD

*University of California, San Diego, CA*: Douglas Conrad, MD; Xavier Soler, MD, PhD; Andrew Yen, MD

*University of Iowa, Iowa City, IA*: Alejandro P. Comellas, MD; Karin F. Hoth, PhD; John Newell, Jr., MD; Brad Thompson, MD

*University of Michigan, Ann Arbor, MI:* MeiLan K. Han, MD MS; Ella Kazerooni, MD MS; Wassim Labaki, MD MS; Craig Galban, PhD; Dharshan Vummidi, MD

*University of Minnesota, Minneapolis, MN*: Joanne Billings, MD; Abbie Begnaud, MD; Tadashi Allen, MD

*University of Pittsburgh, Pittsburgh, PA*: Frank Sciurba, MD; Jessica Bon, MD; Divay Chandra, MD, MSc; Joel Weissfeld, MD, MPH

*University of Texas Health, San Antonio, San Antonio, TX*: Antonio Anzueto, MD; Sandra Adams, MD; Diego Maselli-Caceres, MD; Mario E. Ruiz, MD; Harjinder Singh

### Supplementary Methods

#### Patient and public involvement

This study relied on existing research cohort and biobank data and does not involve any new data collection or direct interaction with the participants. Patients and members of the public were not involved in the design, analysis, or reporting of this work.

#### Additional cohort details

##### COPDGene

COPDGene has three completed study visits approximately 5 years apart. Pulmonary function tests were conducted at baseline enrollment as well as at 5- and 10-year follow-up visits. Genotyping was carried out using the Illumina HumanOmniExpress array, and genotyping for the Z and S alleles of the *SERPINA1* gene (which encodes alpha-1 antitrypsin, a known monogenic cause of COPD) was performed for all participants. Those with severe alpha-1 antitrypsin deficiency were excluded from the study. Imputation was conducted using the Michigan Imputation Server^1^, applying the Haplotype Reference Consortium panel^2^ for white participants and the 1000 Genomes Phase I v3 Cosmopolitan reference panel for African American participants^3^. Variants with an imputation quality score (r²) ≤ 0.3 were excluded from further analysis.

##### ECLIPSE

Longitudinal assessments include spirometry, chest CT imaging, blood sample collection, and exacerbation tracking over a three-year period. Spirometry and exacerbation data were collected every six months during the study. Genotyping was performed using the Illumina HumanHap 550 V3 array with imputation to the Haplotype Reference Consortium panel^4^.

#### Proteomic data

##### COPDGene SomaScan proteomic data

SomaScan 5K v4.0 has been measured on 5,670 participants from frozen plasma from the five-year follow-up study visit; 4,776 proteins were analyzed. Plate hybridization, median signal normalization, and plate scaling and calibration of SOMAmers were performed to control for variability across array signals, inter-run variability, inter-assay variation between analytes and batch differences between plates. Median normalization to a reference using adaptive normalization by maximum likelihood is applied within SOMAmer dilution group to quality control replicates and on individual samples to remove edge effect and technical variance. Data were log2-transformed prior to statistical analysis. Further details regarding preparation of SomaScan data has been previously published^5^.

##### UK Biobank Olink proteomic data

Olink Explore 3072 platform has been collected on 54,219 UK Biobank participants as part of the UK Biobank Pharma Proteomics Project^6^. We included individuals from UK Biobank ≥ 40 years of age with proteomic and spirometry data. We excluded individuals with GOLD 1 COPD or preserved ratio impaired spirometry (PRISm)^7^. To address the class imbalance between COPD cases and controls, we then performed 1:1 propensity score matching on age, sex, and pack-years of smoking using the MatchIt R package^8^.

#### Single-trait polygenic risk scores

PRSs were obtained from the PGS Catalog for C-reactive protein (PGS003527)^9^, venous thromboembolism (PGS003332)^10^, type 2 diabetes (PGS000014)^11^, heart failure (PGS004462)^12^, hypertension (PGS002765)^13^, platelet count (PGS002343)^14^, obstructive sleep apnea (PGS003479)^15^, neutrophil (PGS000105)^16^, anxiety disorder (PGS004451)^12^, cor pulmonale (PGS002049)^17^, major depressive disorder (PGS002789)^18^, allergic respiratory disease (PGS001109)^19^, albumin (PGS002099)^17^, coronary artery disease (PGS000013)^11^, and respiratory infections (PGS000925)^20^.

We also calculated several existing PRSs including for a deep learning model of raw spirometry data (Cosentino et al.^21^), FEV_1_ and FEV_1_/FVC^22^, body-mass index (BMI)^23^, idiopathic pulmonary fibrosis^24^, and asthma^25^. For this study, we also constructed additional PRSs using the lassosum penalized regression method^26^. These included scores for emphysema^27^, peak expiratory flow^28^, eosinophils^29^, and smoking (i.e. cigarettes per day)^30^.

#### Development of a multi-trait polygenic risk score

PRSmix provides a framework to construct polygenic risk scores tailored for complex traits with sub-phenotypic variation^31^. PRSmix+ extends this approach by incorporating phenotype stratification and optimizing score construction through meta-modeling, addressing challenges such as phenotypic heterogeneity and population diversity. By incorporating both primary and secondary phenotypes, PRSmix+ has demonstrated improved performance for multifactorial diseases^31^. This approach is especially suited to COPD, a condition with heterogeneous clinical presentations and phenotypic overlap.

We calculated Pearson correlation coefficients between PRSs (including PRS_multi_) in COPD NHW individuals and performed hierarchical clustering and visualized results with a heatmap to visually assess correlation structure amongst PRSs.

We performed area under the receiver operating characteristic curve (AUC) analyses using the pROC R package^32^. We compared a clinical model (age, sex, pack-years of smoking, BMI, and genetic principal components), PRS_multi_, and the highest weighted individual PRS identified by PRSmix+, and combined models with PRSs and clinical covariates. Statistical comparisons between models were performed using DeLong’s test, with p-values < 0.05 considered significant.

#### Meta-analysis

As an additional sensitivity analysis, we conducted a leave-one-out meta-analysis excluding COPDGene NHW, since this cohort was used to develop the PRS_multi_ weights and we aimed to ensure overfitting was not the primary driver of observed effects.

#### Drug repurposing analysis

For example, a protein meeting criterion for both FEV₁ and FEV₁/FVC would be ranked higher than one associated with only a single trait. Each protein was cross-referenced with the OpenTargets database to identify corresponding druggable targets^33^, existing drugs, and ongoing or completed clinical trials. We also identified proteins in which the association of each protein level with COPD affection status, frequent exacerbations (≥ 2 exacerbations/year vs. no exacerbations), and highest vs. lowest quintile of the PRS_multi_ in COPDGene NHW participants was consistent across all three outcomes.

### Supplementary Tables

See SupplementaryTablesPRSmix_Zhang_Konigsberg_07032025.xlsx.

### Supplementary Figures

**Supplementary Figure 1.** Heatmap showing hierarchical clustering of Pearson correlation coefficients between PRSs input into PRSmix+.

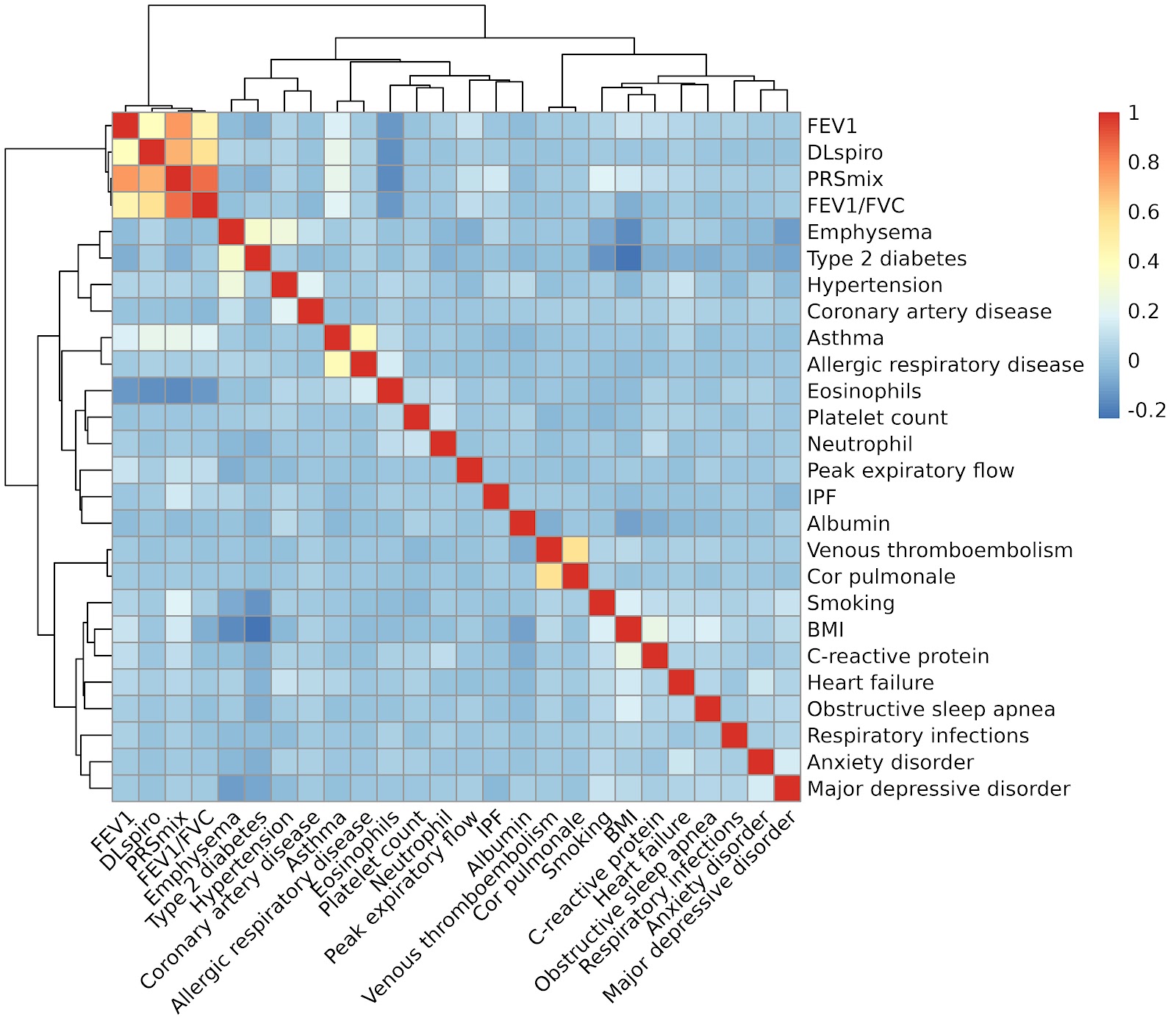

**Supplementary Figure 2.** Forest plot showing the random effects meta-analysis of multivariable negative binomial associations of PRS_BMI_ on exacerbations.

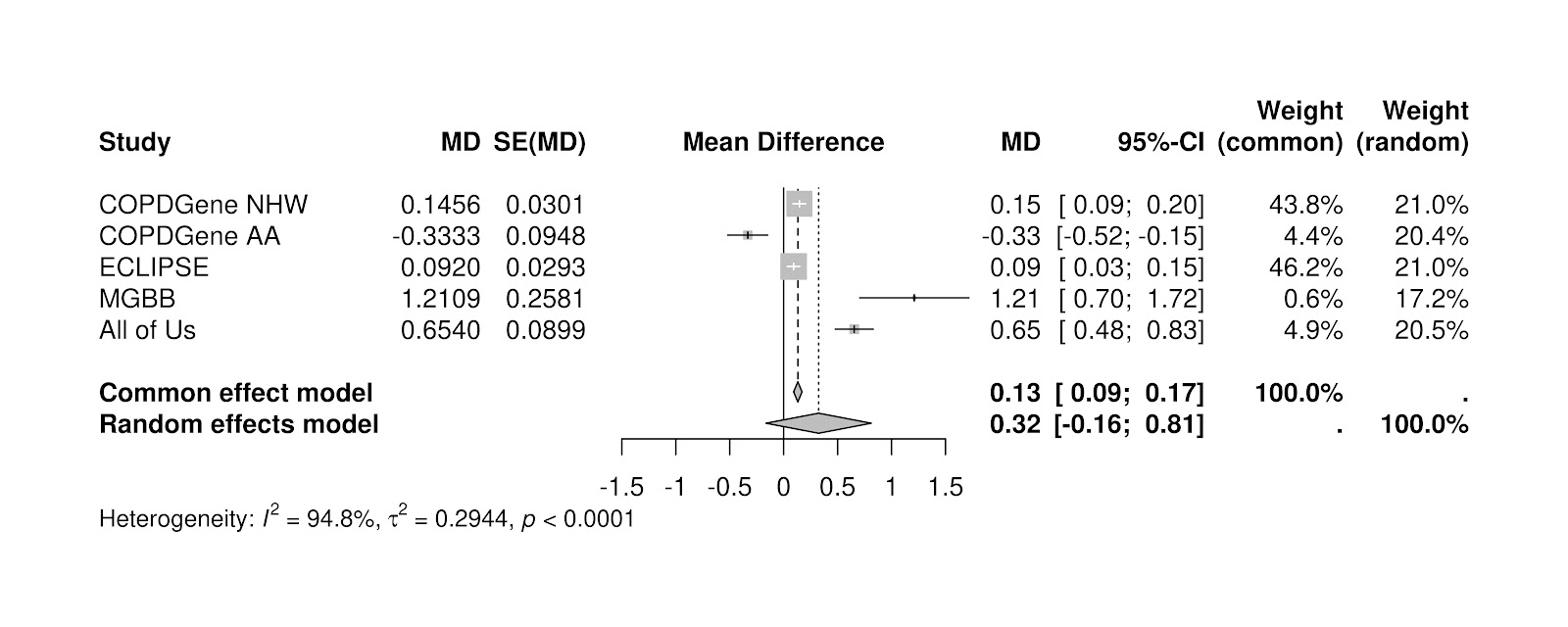

**Supplementary Figure 3.** **Funnel plots of meta-analysis results for COPD and exacerbations.** Funnel plots displaying meta-analysis results for (A) COPD and (B) COPD exacerbations.

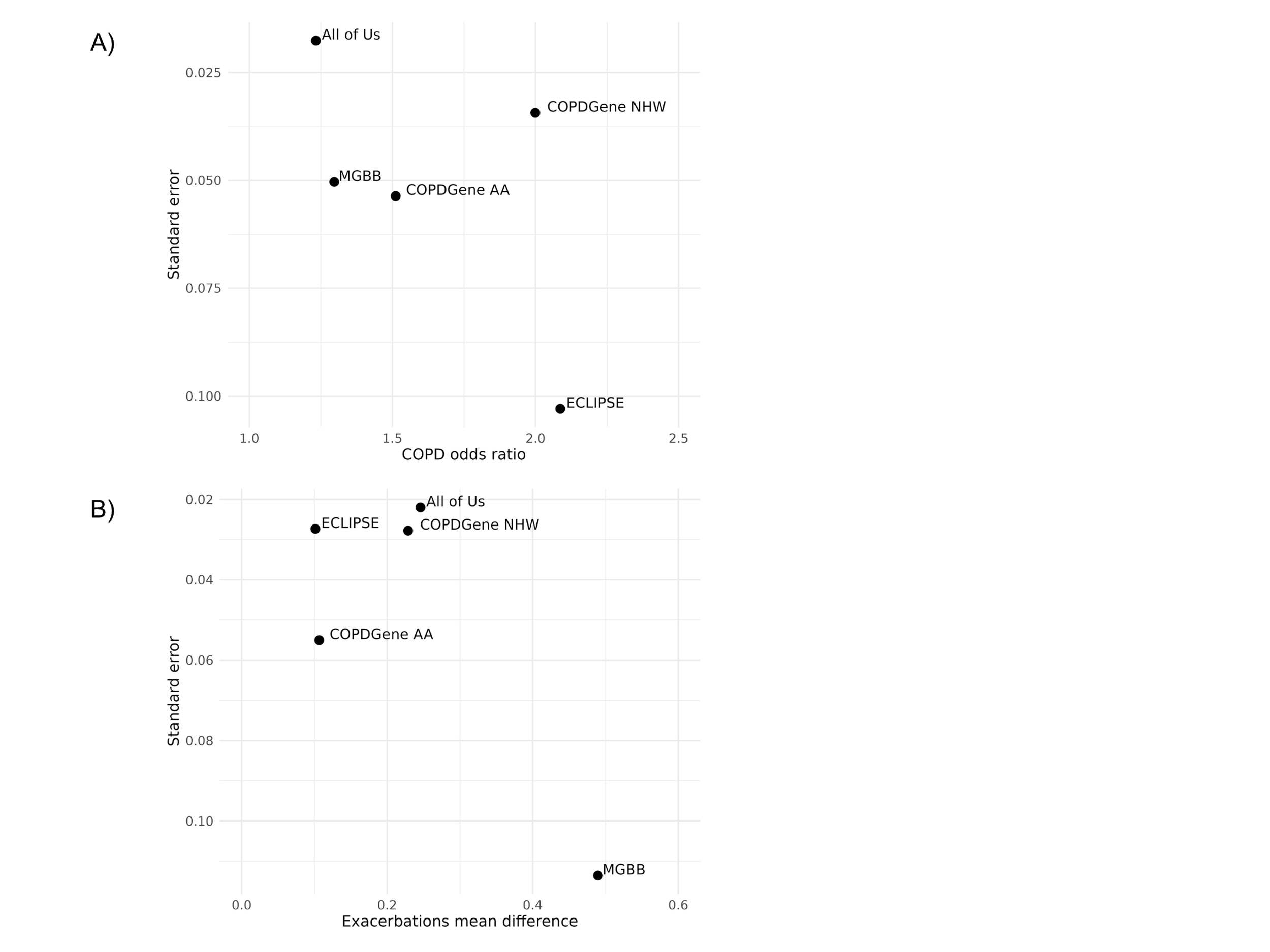

**Supplementary Figure 4. Leave-one-out meta-analysis for COPD (A) and exacerbations (B).** Meta-analyses excluding COPDGene NHW participants were performed as this cohort was used to tune PRS_multi_ weights and could be prone to overfitting.

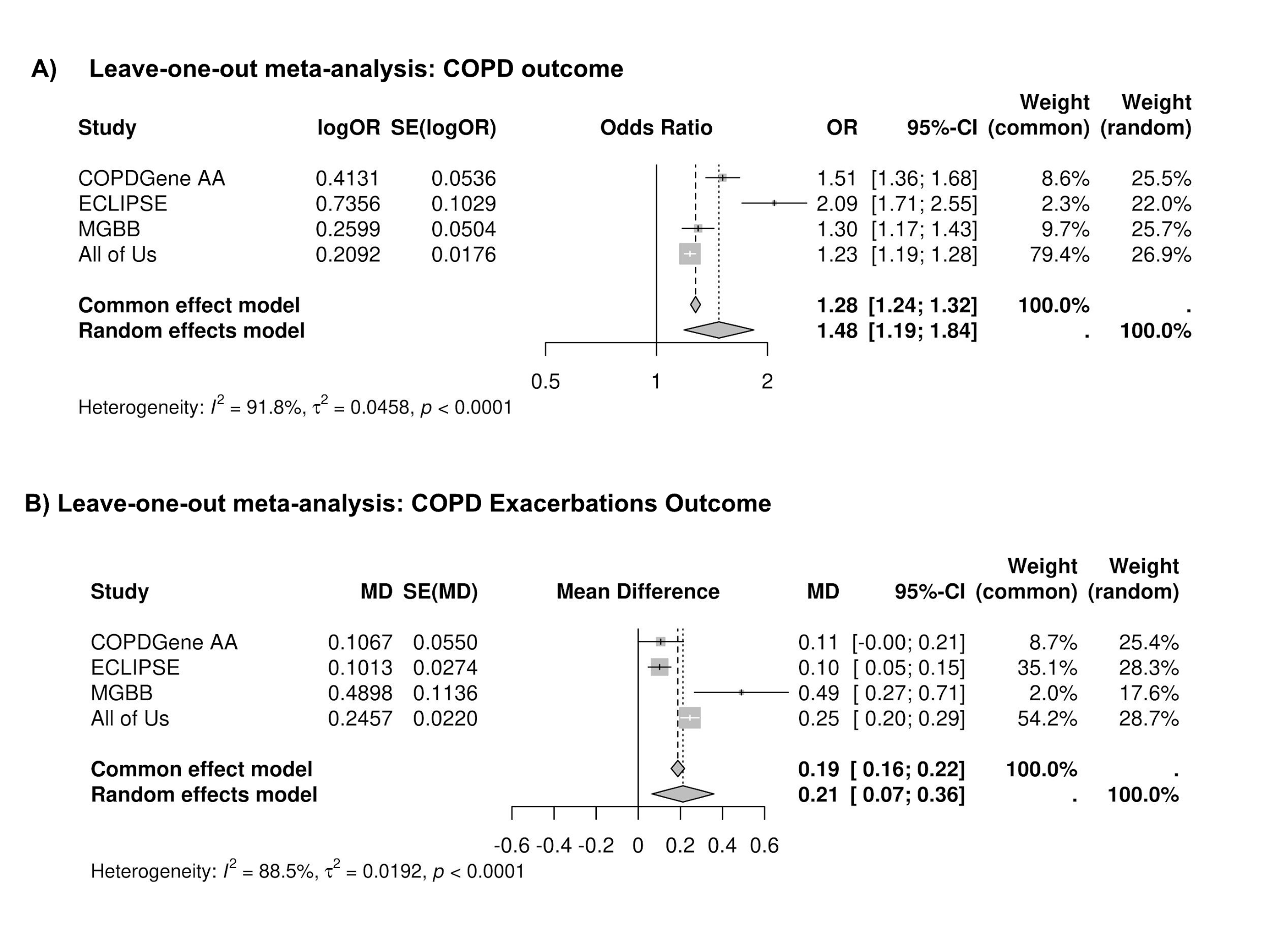

**Supplementary Figure 5. ROC curves for PRS-based models of COPD and exacerbations.**
Receiver operating characteristic (ROC) curves for models predicting (A) COPD and (B) exacerbations. Models include combinations of clinical covariates, PRS_multi_, PRS_ratio_, and genetic principal components.

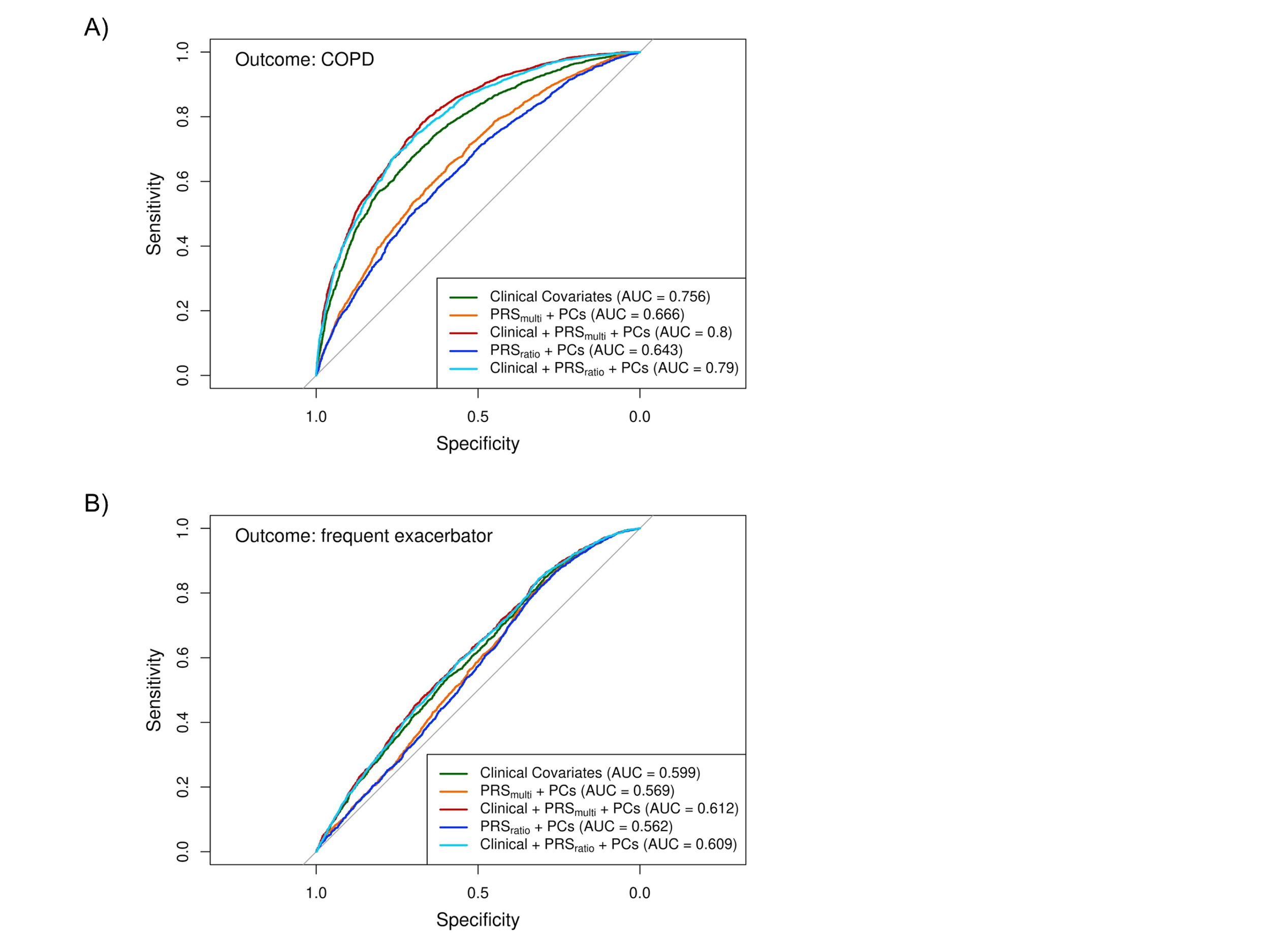

**Supplementary Figure 6. Protein Associations with COPD Outcomes.** Of the 73 genetically-predicted proteins identified to be associated with COPD exacerbations, eight proteins were associated with COPD affection status (left), frequent exacerbations (≥ 2 exacerbations/year vs. no exacerbations) (middle), and significantly differentially expressed between the top and bottom quintile of the PRSmulti (right) in COPDGene NHW participants. Boxplots for each association with interquartile ranges are displayed. P-values for Student t-tests are also displayed.

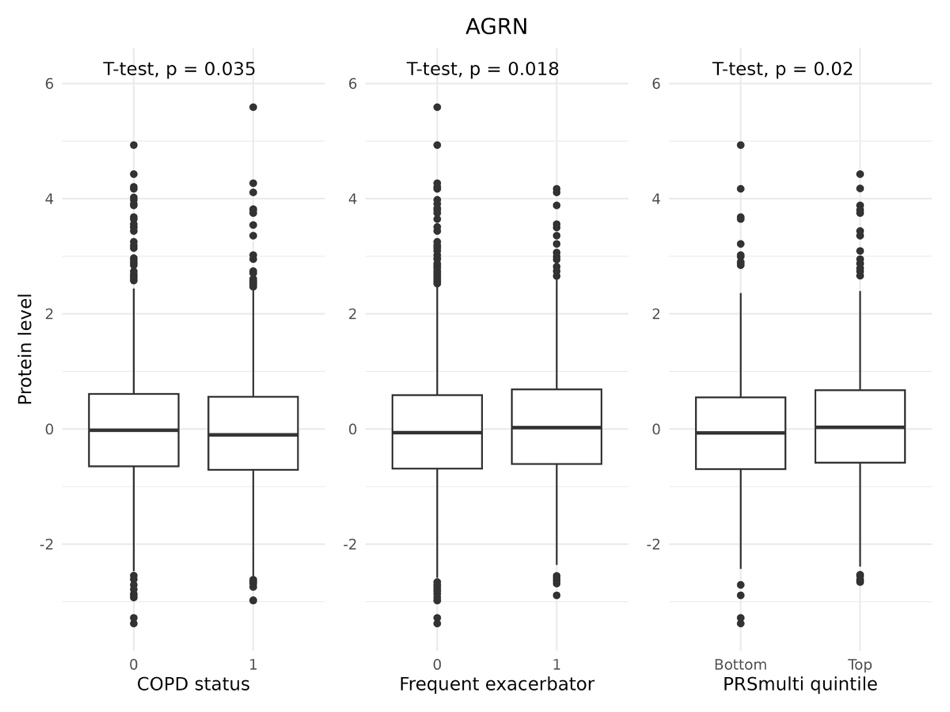

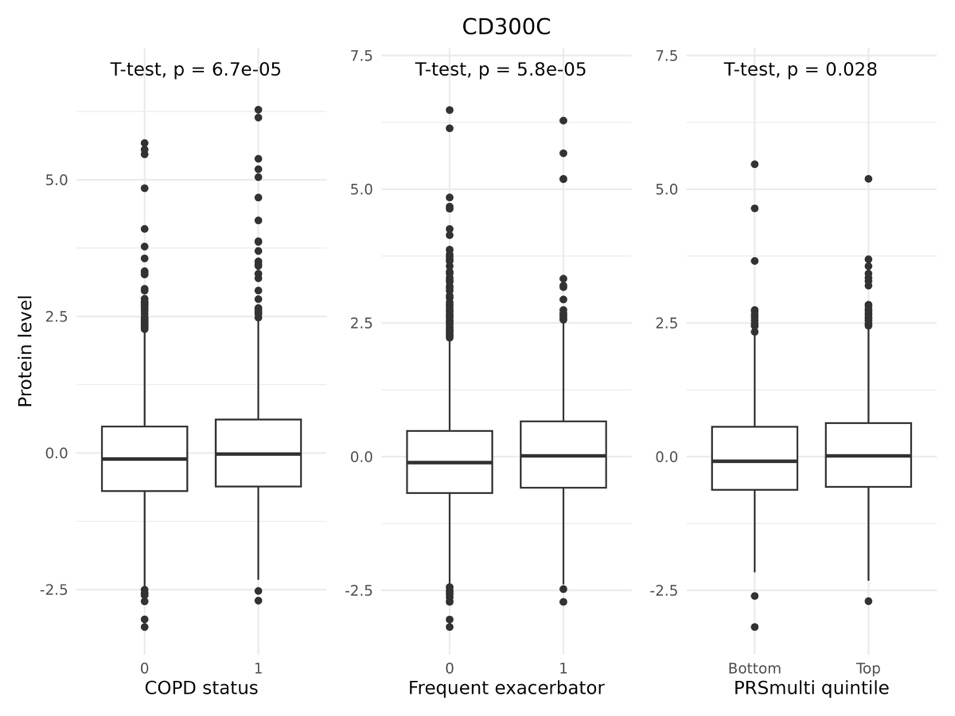

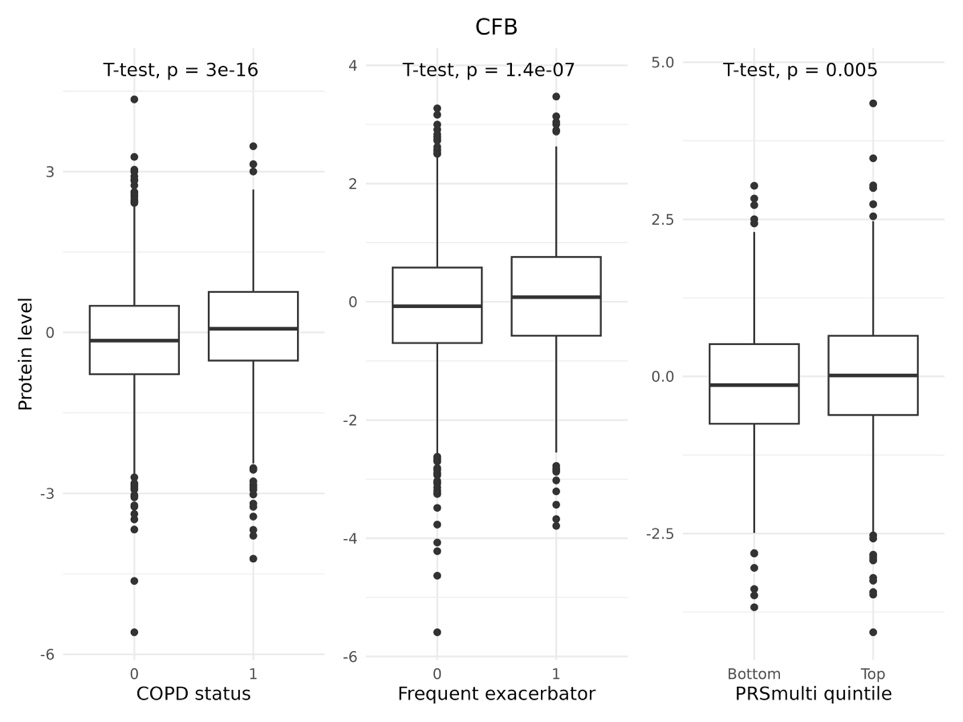

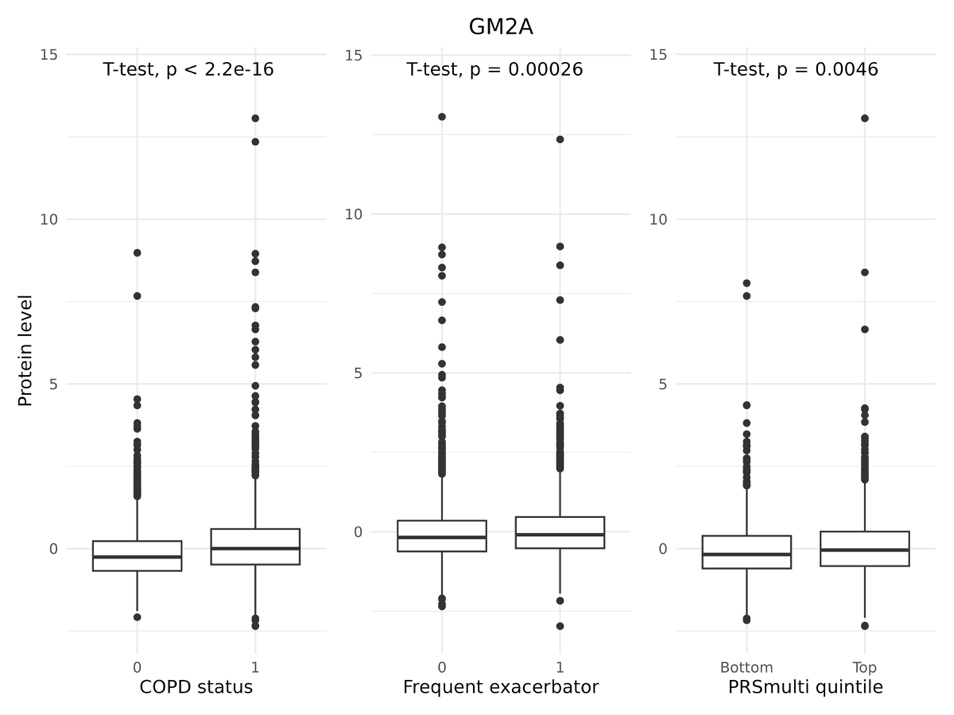

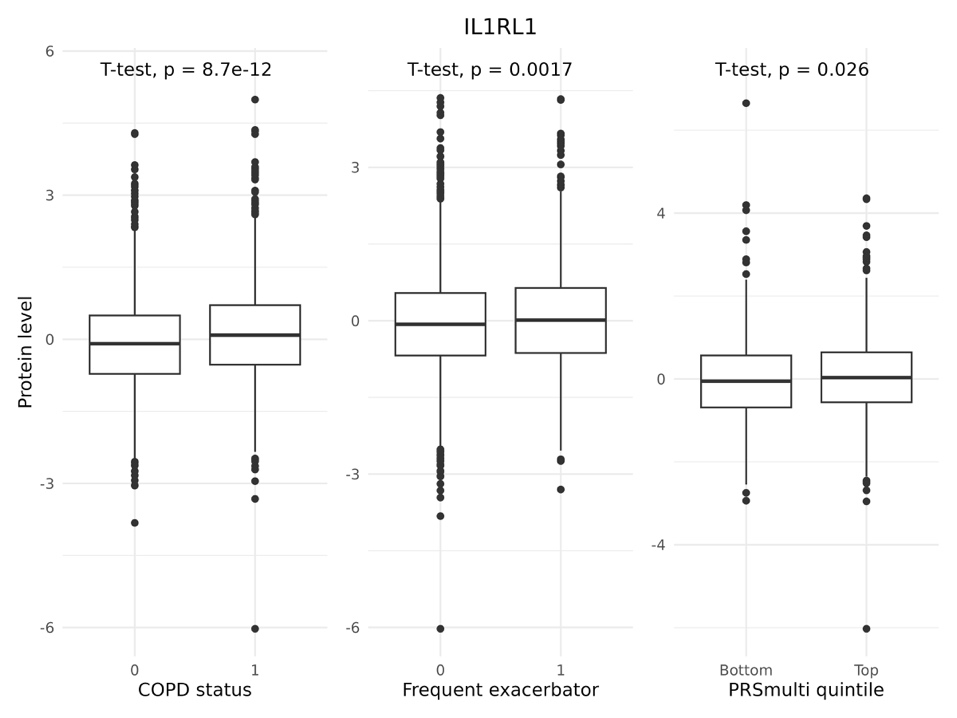

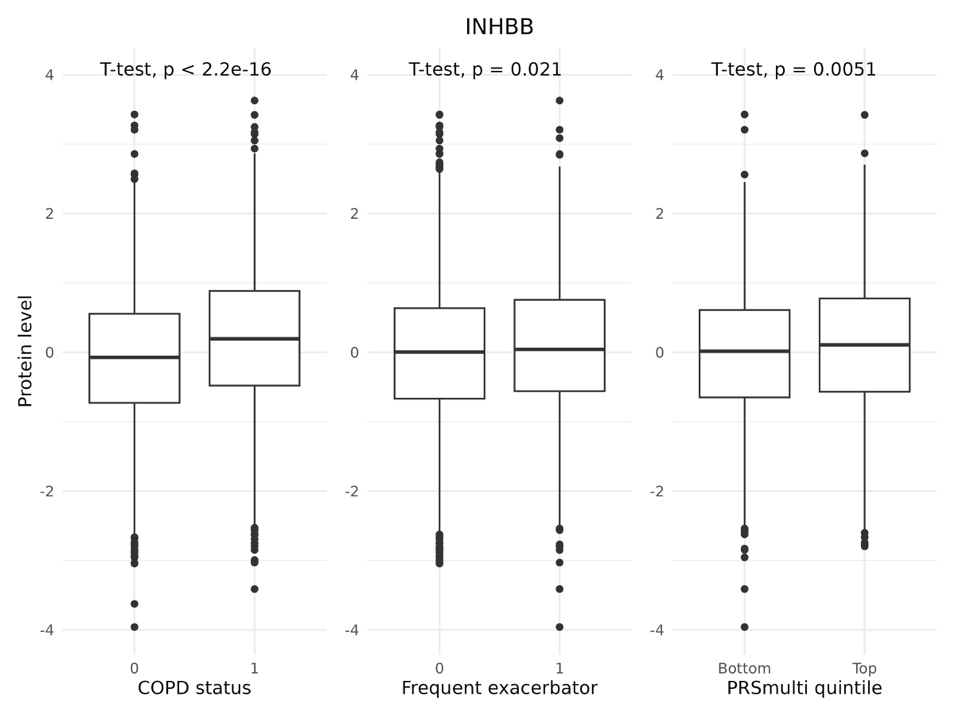

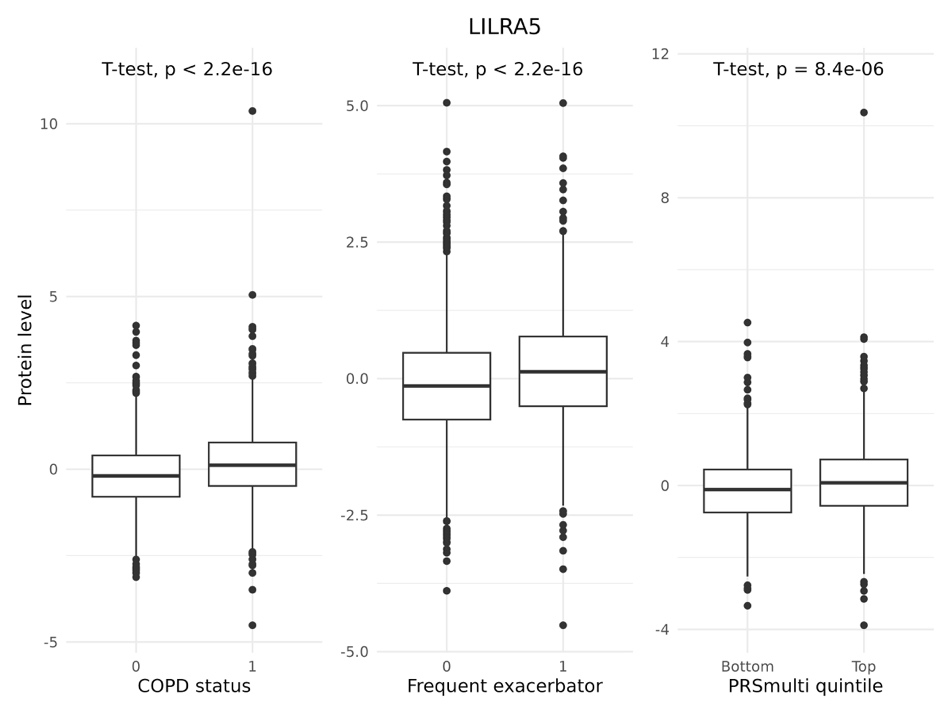

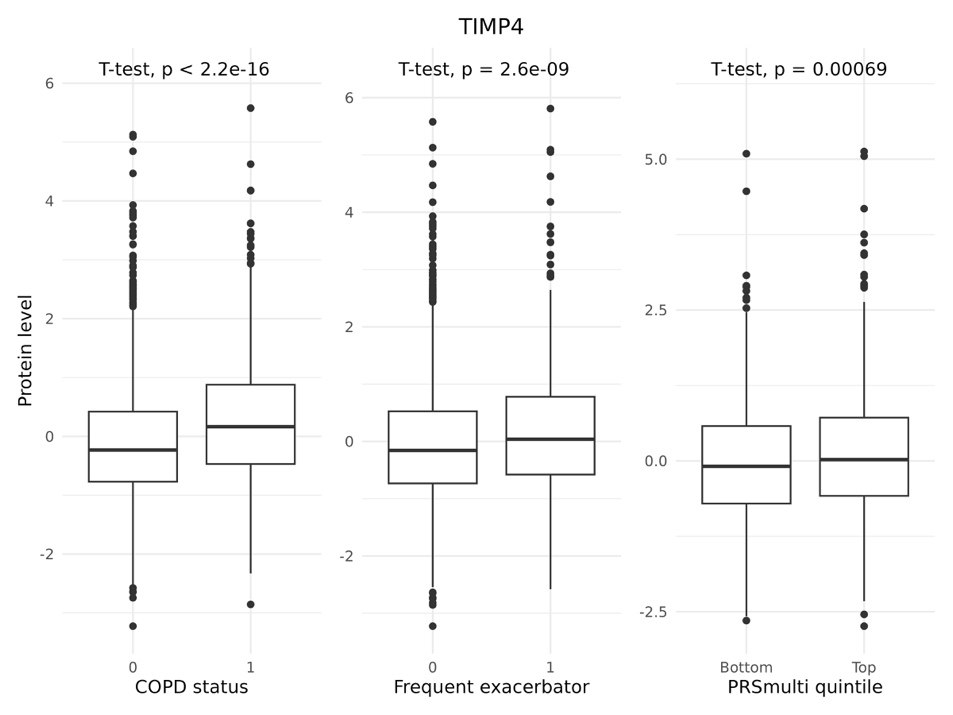

### Supplementary Discussion

Previously, we showed that individuals with low BMI disproportionate to their genetically-predicted BMI (e.g., COPD patients with cachexia) have higher mortality risk^23^. In the current study, PRS_BMI_ was strongly associated with exacerbations in biobank cohorts, though it did not reach significance in meta-analyses, largely due to a flipped effect in COPDGene African Americans. Whether these discrepancies reflect differences in COPD definitions, ascertainment, or cross ancestry genetic prediction remains unclear. Given the links between BMI, mortality, and exacerbations, further investigation is needed.

The inclusion of PRS_IPF_ in PRS_multi_ is consistent with prior evidence that IPF and COPD share overlapping and distinct genetic loci^34^. Smoking and C-reactive protein, both associated with COPD progression and inflammation, explain their inclusion in PRS_multi_^35-37^.

This study builds upon prior work in several key ways. Unlike our prior work on PRSs for COPD, which focused on just two spirometry traits without incorporating negative binomial modeling for exacerbations or expanding analyses to biobanks, our work addresses these gaps. He et al. applied a multi-trait GWAS framework that included genetic associations for asthma, COPD, lung cancer, smoking, and spirometry traits^38^; we significantly expanded this list including 25 traits based on clinician input and literature review of COPD comorbidities, risk factors for exacerbations, and existing biomarkers. We then utilized PRSmix+ to identify relevant traits. In one of these two prior studies, either spirometry-defined COPD in research cohorts or ICD-defined COPD in biobank cohorts were tested, but we tested both definitions together and observed robust but highly heterogeneous associations across cohorts. Finally, we placed a stronger emphasis on exacerbations, carefully defining our phenotype and utilized more advanced negative binomial models compared to simple linear or logistic regression models. Taken together, our study provides a more robust foundation for understanding the shared genetic architecture of COPD and the relationship to exacerbations.
